# eHealth Treatment Recommendations for the Common Cold across 13 European Countries: Correlation with Antibiotic Use

**DOI:** 10.1101/2025.10.06.25337441

**Authors:** Suzy Huijghebaert, Koenraad Tilkin-Franssens, Walter Geerts

## Abstract

**Background:** Antibiotic consumption (ABC) often ensues from patients’ misbelief that ABs are needed to treat the common cold. To test the impact of eHealth communication, the treatment recommendations on eHealth portals for patients/consumers were explored, modelled and correlated with ABC.

**Methods:** Cross-sectional study: identification of eHealth “common colds” pages (13 European countries); qualitative review of the treatment recommendations for antibiotics, nasal saline drops/irrigation, decongestants, other medicines; conversion to scores and correlation with ABC (ECDC-community sector 2022). Traditional use of saline was taken into account as co-variable.

**Results:** e-sites generally explained why ABs are not needed, yet this message and position, as well as treatment e-recommendations highly differed. ABC differed by major treatment e-recommendations (*p* = .017), while a strong negative correlation was found between ABC and the treatment recommendation scores, indicating that ABC was lowest, if the eHealth site recommended saline first-line, while ABC increased if rather decongestants (no saline) were proposed, and even more, if emphasis on various medicines and complications (Spearman *r*_s_ = -.945, *p* < .001).

**Conclusions:** The strong inverse correlation of ABC with SNI/treatment recommendations suggests that eHealth recommendations for the common cold may significantly affect ABC, and thus the patients’ need for ABs. The findings are in line with independent observations that the messages on prudent use of ABs need empowerment by offering patients effective symptom control. eHealth messages and solutions proposed need urgent further study as to change patient’s expectations and reduce ABC.

**Key points:**

- The common cold is a major driver of antibiotic consumption (ABC) in the community.
- Analysis of the common cold pages of 13 European public eHealth sites revealed they all discouraged ABs but the messages and position on the e-page highly differed.
- The highly variable ABC strongly inversely correlated with recommendations for nasal saline (traditional use as co-variable).
- ABC was lowest, if the eHealth site recommended nasal saline, yet increased if decongestants (no saline) first line, and even further if communicating many medicine options or complications.
- Findings confirmed that messages on prudent use of ABs need empowerment by offering an accessible effective alternative for symptom control.
- Education of the public and physicians, and well-structured, persuasive public eHealth communication about the common cold, recommending nasal saline, may help to reduce ABC. Further research is warranted.

## Introduction

The development of antimicrobial resistance (AMR) against bacterial pathogens has been attributed to the widespread use of antibiotics (ABs): AMR has been directly associated with worse clinical outcomes, such as longer hospital stays and excess mortality, so that from a global public health perspective, AMR counteracts the United Nations Sustainable Development Goals 2015 [1]. Also, the World Health Organization (WHO) and European Commission have set priorities to address AMR, having stimulated multiple programmes to combat AMR: many of these focus on education as to reduce antibiotic consumption (ABC) [2,3]. The role of healthcare professionals has thereby been recognised as “gate-keepers” for appropriate use and for educating the patients [1]. Also, the role of eHealth tools to support clinical decisions has increasingly been acknowledged [4]. However, little attention, if any, has been paid on the role of public eHealth treatment recommendations, as to guide the patients’ attitudes, when it comes to the treatment of common complaints, such as the common cold and upper respiratory tract infections (URTIs). This aspect, though, needs attention for several reasons, such as the increasing trend in selfcare through consultation of the Internet, and the impact of e-guidelines on artificial intelligence (AI)-generated answers to health-related questions. Moreover, there is an increasing need for models to reflect the multidimensional, dynamic, and relational nature of eHealth recommendations with patients’ and clinicians’ practices [5]. This prospective study therefore aimed at identifying the public eHealth recommendations for the common cold across Europe, for saline nasal irrigation/spray/drops (SNI) in particular, and at developing and testing a model to evaluate whether e-treatment recommendations may affect ABC. We followed thereby the following rationale.

Firstly, ABC strongly varies across Europe, despite the European’s Council recommendation on the prudent use of antimicrobial agents in human medicine since 2001 [6], and despite many longstanding local Health policies and multifaceted campaigns, raising the awareness about AMR and emphasizing the inefficacy of ABs for respiratory viral infection: there impact has often been found limited or insufficient in reducing ABC [7,8,9,10]. A recent systematic review revealed both increases and decreases in ABC, with often replacement of one AB by another AB [11]. On the other hand, inappropriate treatment of acute URTIs seem to be a strong driver of AB prescribing in primary care [e.g. 12,13,14,15]. Despite the earlier and more recent global strategy set by the European Council (One Health programme) across Europe [6,16,17], the hopeful reduction of ABC in the community, observed during the two COVID-19 pandemic years 2020-2021, was countered by a rebound in 2022 [8]. This was attributed to resurgences in URTIs after 2 years of restricted mobility and social contacts during the COVID-19 pandemic [8]. Rebound also occurred despite the proposition of a more reserved policy for prescribing AB in primary care [18]. Public eHealth recommendations may determine the first steps of patients in their decision to self-medicate with, or to seek a doctor for, an AB.

Secondly, normal (isotonic) SNI and saline gargling are simple, inexpensive nonpharmacological hygiene interventions, traditionally used in several European countries to treat the common cold and accessible to everyone. SNI was already proposed in 2003 as “*a simple, inexpensive treatment that relieves the symptoms … and could help minimize AMR*”, because patients treated with SNI rely less on other medications and make fewer visits to physicians [19]. Apart from its appreciated efficacy to treat URTIs in routine clinical practice in children [20], this proposition in adults is supported by earlier studies [21,22,23] and a recent large, randomised, controlled study in UK primary care practices, enrolling 13799 patients with least one comorbidity or risk factor associated with respiratory illness: SNI reduced overall illness duration of URTIs, the number of days with more severe symptoms and the number of lost days of work or normal activities; there was also a modest reduction in the ABC, when compared with usual care [24]. Therefore, SNI was selected in this study as first-line e-recommendation among remedies that patients can apply themselves to relieve the common cold, and so may affect patients’ need for an AB.

Thirdly, eHealth recommendations are likely to affect the patients’ attitudes to ABs: from a communication point of view, an official public eHealth site typically follows a “knowledge deficit model” [25]. The authorities provide “knowledge” or information on, and recommendations for (self-)treatment: they thereby aim at educating patients/readers about the disease and prudent use of AB, with the goal of helping out and creating correct “attitudes” to treatment, ultimately expected to lead to lower ABC in “practice”. Yet, misbeliefs about the usefulness of ABs and self-medication with ABs are common in URTIs [26,27,28]. There may be considerable pressure of patients on the physicians for prescribing an AB [29]. On the other hand, a study revealed that parents appreciate information and reassurance from the doctor, considering prescription of medication less important [30]. Appropriate counseling resulted in a small reduction in ABC compared to usual care [24]. Therefore, also the messages on ABs on the e-pages of the common cold were identified in this study. As vice versa, it is also known that extensive information/education can lead to the contrary if the communication creates uncertainty [25], the study also evaluated whether there was considerable focus on other medications and/or complications on the e-page. At least in pharma communication on the Internet, the principle of disease mongering is a well-known practice, whereby education of patients, and linking ordinary ailments to more serious medical problems, aim at driving patients to seek a doctor, so to maximise opportunities for prescribing [31]. Unwillingly, this may be an overlooked aspect in public eHealth education. These considerations led to a theoretical framework proposition (**Figure S1)**, allowing to work out a model aiming at comparing the public treatment e-recommendations for the common cold across 13 European countries, with focus on SNI or other options as first-line remedies, and at correlating the outcomes with the highly variable ABC across Europe. Our theoretical framework also integrated the traditional use of SNI, as co-variable per country: this is important, as the correlates of self-medication practices and online consultations have revealed that the ultimate adoption (or non-adoption) of the e-recommendations is rather defined by the off-line (thus traditional) “habits” [26,32].

To note: this prospective study did not investigate the patients’ attitudes itself following the exposure to the e-site, or the doctor’s AB prescribing, but rather aimed at a prospective translational approach whereby input (communication & clinical recommendations) is correlated with the output – the ABC prescribed in the clinicians’ practice. Further, in this article, “nonpharmacological” interventions refer to the use of traditional selfcare remedies, but not to the nonpharmacological hygiene recommendations, that were imposed since the COVID-19 pandemic, such as washing hands, cough hygiene and social distancing (systematically implemented on the eHealth sites). Moreover, as eHealth encompasses many different aspects, such as “technologies” for finetuning treatment and guiding prescribers towards more correct AB use in healthcare [33], it is to note that eHealth communication in this article only refers to the public communications, typically found on patient portals, aiming at providing health education and guidance to patients for selfcare. Our investigation fits the WHO action plan calling for “action to understand accelerators and overcoming barriers” [34], however rather focusing on the eHealth communications reaching consumers or patients seeking advice for the common cold. The results are discussed extensively with reference to the existing literature on attitudes to and use of ABs, and needs for further study identified.

## Materials and Methods

### Inventory of Recommendations

Authorities’ official eHealth site(s) were searched for and screened, or - if not readily identified or accessible/screenable as for a consumer - other eHealth sites with recommendations, sequentially as popping up on the Internet per individual country [Google searches between July 2023 and June 2024 (verifications up to 10.08.2025)]. As a guide for selecting the countries, the ECDC list was used as published by Ventura-Gabarró *et al.*, 2022) [8] and reasons for exclusion were listed (**Table S1**). Countries were excluded for the following reasons: (1) no reliable e-site identified, too little information or inadequate translations (e.g., Slovenia, Bulgaria, Greece, Portugal); (2) no information on defined daily doses (DDDs) in the ECDC list (United Kingdom); (3) (touristic) islands that may have different dynamics in ABC (e.g., Cyprus); (4) countries likely relying on other countries’ e-sites (e.g., Ireland consulting UK NHS); (5) countries with high ABC attributed to exceptional use in a particular indication other than URTI (Poland): for motivation per country, see **Table S1**. This allowed to select 13 European countries: all (except maybe Estonia) are known having initiated national programmes to reduce ABC in the past two decades [17]. Typical target words in the local language (**Table 1**) were linked to the country’s internet domain extension. Texts of the common cold e-page(s) were retrieved and translated, and with the e-page’s structure listed in an inventory, as to allow easy reidentification, verification and reproducibility of the information analysed. **Table 1** lists the target words used, the countries’ internet extensions and relevant hits (= eHealth sites used for the qualitative evaluation). The links and translated texts, and major structure of the common cold pages on the e-site are included in the inventory in the Supplement **Table S2**.

**Table 1.** The target words used and official or main eHealth portal by the health authorities, or if unavailable, by representative medical organisation(s)(Ext. = Internet domain extension). For more details, see inventory in Table S2.

| Country<br>(Ext.)<br>Health Authority | Target word<br>Common cold<br>(local language) | First hit (or if not relevant, the next alternative sites) |
| --- | --- | --- |
| **Austria**<br>(.at)<br>Gesundheitsportal | Erkältung | **First hit:** Netdoktor.at: recommending inhalations first-line (specialist medical texts, belonging to Burda Verlag)<br>**Second hit:** [Gesundheit.gv.at](https://www.gesundheit.gv.at)<br>= Öffentliches Gesundheitsportal Österreichs<br>= Hosted by Bundesministerium, Gesundheit, Pflege, and Konsumentenschutz → for treatment, the link goes to the 'Österreichische Sozialversicherung':<br>https://www.sozialversicherung.at/cdscontent/?contentid=10007.862784&portal=svporta. |
| **Belgium**<br>(.be)<br>(Flanders)<br>(gezondheids)overheid | verkoudheid | **None** found among the first 10 hits.<br>Apart from Factchecker, Sciensano (= epidemiology reporting), and a university, all sites appeared to be sponsored by press, pharma industry or hospitals.<br>Also, Dutch health sites popped up. |
| (Wallonia)<br>autorité de santé | rhume | **None** of the sites were directly practical; information was fragmented with a focus on communication on decongestants on the internet sites:<br>CBIP (Centre Belge d'Information Pharmacothérapeutiques).<br>AFMPS: Agence Fédérale de Médicaments et des Produits de Santé. |
| **Denmark**<br>(.dk)<br>Sundhedsmyndighed | forkølelse | [Sunhed.dk](https://www.sunhed.dk)<br>= the official Danish eHealth Portal for the public Danish healthcare services. |
| **Estonia**<br>(.ee)<br>Tervishoiuasutus | nohu | **First hit:** [Kroonilinenohu.ee](https://www.kroonilinenohu.ee)<br>= free information source on health but unclear who hosts the site; the hit rather referred to 'chronic rhinitis'.<br>**First following hits:** [Arst.ee](https://www.arst.ee)<br>= medical website "that connects patients and doctors and introduces health products".<br>https://rakveretervisekeskus.ee/storage/app/media/hingamisteedea53trykk-1.pdf = brochure by Estonian Society of Infectious Diseases, Ambulance Association, Estonian |
|  |  | Association of Paediatricians, Association of Family Nurses of the Estonian Nurses' Union. |
| **Finland**<br>(.fi)<br>terveysviranomaiset | flunssa | **First hit:** Duodecim Terveyskirjasto<br>= the site hosted by the Finnish Institute for Health and Welfare |
| **France**<br>(.fr)<br>autorité de santé | rhume | **First hits:**<br>Official agency : ANSM (Agence Nationale de Sécurité du Médicament et des produits de santé)<br>Santé Publique France (République Française): covering rather articles on disease topics<br>https://www.santepubliquefrance.fr<br>**After adding target word 'antibiotics':**<br>**Santé.fr:**<br>= official French eHealth site, also rather publishes on selected topics.<br>**Other e-sites** screened, initially popping up earlier, and ranking higher for e-visits: VIDAL, TopSanté. |
| **Germany**<br>(.de)<br>Gesundheitsportal<br>Gesundheitsbehörde | Erkältung | **First hit:** Gesundheitsinformation.de<br>https://www.gesundheitsinformation.de/ueber-uns/das-iqwig/<br>= hosted by Das Institut für Qualität und Wirtschaftlichkeit im Gesundheitswesen (IQWiG) |
| **Italy**<br>(.it)<br>autorità sanitaria | raffreddore | **None** found among the first 10 hits. At No. 13 came up the official site: https://www.issalute.it/<br>= Portal by Istituto Superiore di Sanita (ISS)<br>= hosted by Italian National Institute of Health (ISS), the main centre for research |
| **Luxembourg**<br>(.lu)<br>Gesondheet Autoritéit | Schnappf<br>(allgemeng<br>Erkältung) | **First hit:** Santé.lu<br>= hosted by Direction de la Santé |
| **Netherlands**<br>(.nl)<br>overheid gezondheid | verkoudheid | **First hit:** Thuisarts.nl<br>= official eHealth site of the Dutch Health authorities |
| **Norway**<br>(.no)<br>Helsemyndighet | forkjølelse | **Among first hits:** Helsenorge.no (5<sup>th</sup> site)<br>= official eHealth site of the Norwegian Health authorities |
| **Spain**<br>(.es)<br>autoridad sanitaria | resfriado | **Among first hits:** Sanidad.gob.es : one is automatically deviated to "flue" on the portal of the Ministerio de Sanidad y Política Social.<br>https://informaciongripea.sanidad.gob.es/preguntas-y-respuestas-mas-frecuentes.html<br>Sanidad.gob.es is the official public eHealth site by Spanish Ministry of Health (MISAN), yet not to confound with Sanidad.es (unclear who hosts; see Inventory in Supplement for more details) |
| **Sweden**<br>(.se)<br>hälsovårdsmyndigheten | förkylning | **First hit:** Helpline 1147.se<br>= official site with information and services in health and care by the Swedish authorities |

### Qualitative Analysis

The Common Cold page was qualitatively analysed for (1) the presence of a message discouraging or limiting the use of ABs in the common cold and (2) the treatment recommendations: (a) nasal “saline” as part of remedies for selfcare; (b) decongestants; (c) other cold medicines such as antivirals or vaccines. (see also Assumptions). These parameters were noted in colours in the left column of the inventory (**Table S2**), as to allow visualization of their prioritization per eHealth site. To take the traditional use of remedies, such as SNI, into account, countries were screened for traditional use of SNI (nasal drops, spray or irrigation) and gargling, based on internet screening, literature and personal contacts. The findings of the qualitative analysis were summarized per country in the Inventory.

### Antibiotic Consumption: Dataset & Parameters

ECDC publishes annual epidemiological reports on ABC in the EU/EEA: the data set of 2022 was used, more precisely, the community sector consumption of antibacterials for systemic use (anatomical therapeutic chemical [ATC] group J01), quantified as DDD per 1,000 inhabitants per day [8]. In the primary analysis, the class ATC J01 data were used for correlation with the scores and calculation of the mean and median ABC; in the secondary analyses, the datasets from ATC J01C and J01F were used, being the AB subclasses most relevant to the treatment of (U)RTIs. The concept thereby is that ABC in the community can be used as a quantitative surrogate or proxy to reflect the patients’ (attitudes to the) need for an AB in the common cold, for which ABs are easily being misused and also overprescribed: a pivotal role of the patient’s request for an AB on the doctors’ prescribing of ABs has recently been evidenced by a study across 18 European countries [15]. Hence, it can be assumed that ABC is guided by the consequences of self-treatment, and in the case of failure of self-help, by seeking the doctors’ advice for an AB prescription.

### Model, Assumptions & Algorithm

This pilot study used a simplified, straightforward theoretical framework (**Figure S1**), allowing conversion of the qualitative findings into scores and categorizing the countries by scores. This is a common approach in pharma for exploring a potential impact of messages on the internet and their correlation with clinical or economic output. It allows the integration of qualitative data that represent a direct encounter of the internet user (and thus likely his uses/experiences) into quantitative consumption data, so can reveal nuances in usability, missed otherwise by coding quantitative survey outcomes; the resulting category or score allows to prompt further questions for usability improvement [35]. Therefore, to assess the correlation between eHealth recommendations for the common cold and the patients’ attitudes to a need for an AB, the model was constructed whereby (1) the ECDC ABC in the community was considered to largely reflect the patients’ attitudes to a need for an AB (either by self-medication or visiting a physician to receive a prescription); and (2) the recommendations on the eHealth platform were given a score, taking into account the first-line positions of SNI, decongestants/cold medicines, and other (antiviral) medicine, as well as communication on vaccines and complications.

For the assumptions used in the model, see **Figure 1**. The model assumed that, if no effective remedies are convincingly proposed on the eHealth sites, this will lead to a higher ABC, because otherwise, if there is no relief or resolution of the symptoms, the misbelief or conviction remains that an AB is needed to get cured. Proposing decongestants or other medicines first-line (not reducing ABC), rather than SNI (reducing ABC), was assumed to drive patients to buy these medicines, and if not working properly or within the recommended time-limit proposed for use, to drive the patient to seek a doctor for an AB prescription. The model also assumes – in line with the known effects of disease mongering [31] - that messages raising the perception that a medicine is needed, or elaborating on complications as a problem, may faster create the patient’s need for an AB. **Figure 2** summarizes the structured algorithm followed in this pilot study, matching the assumptions.

**Figure 1.**
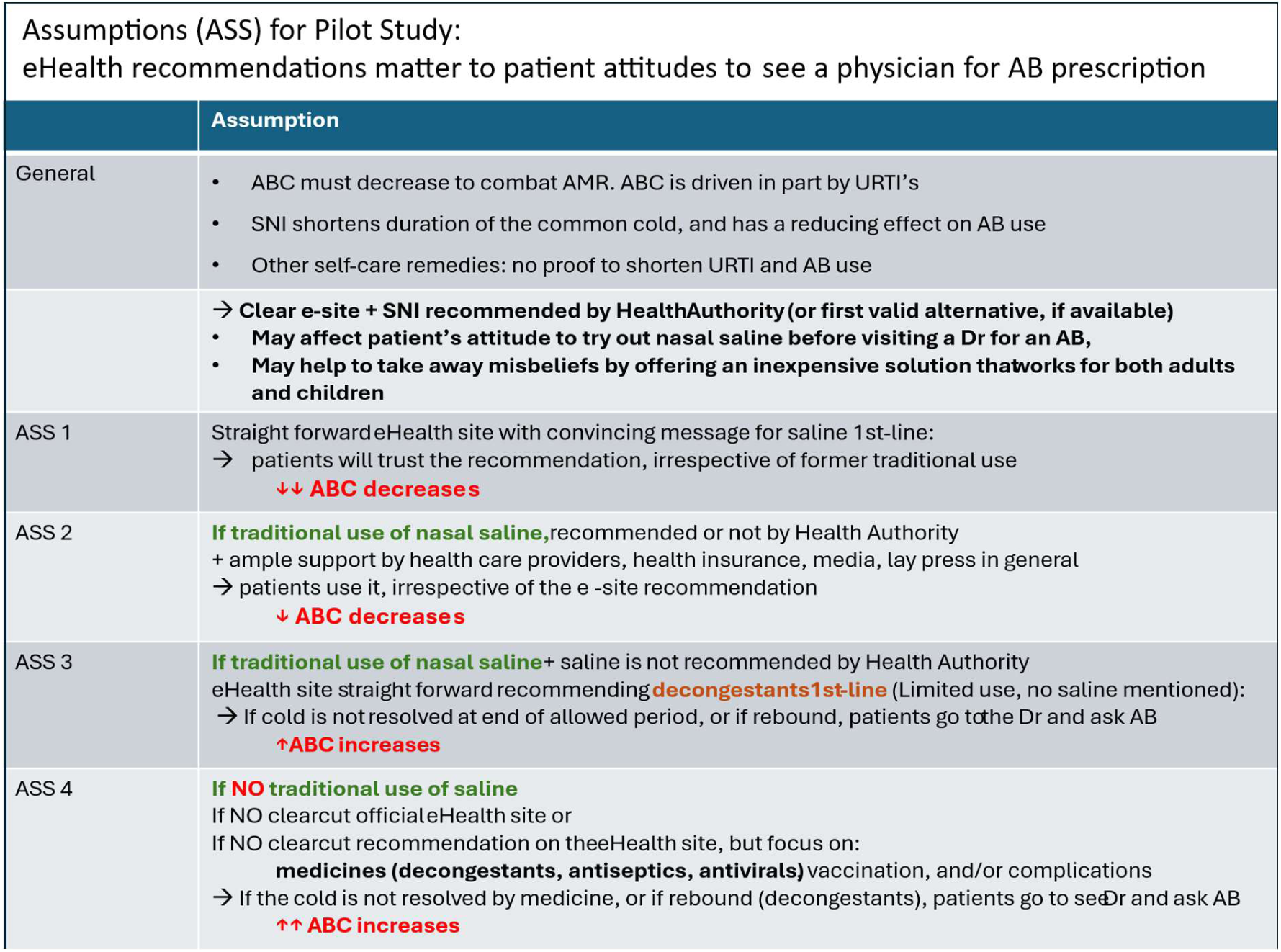
Assumption used for developing the model

**Figure 2.**
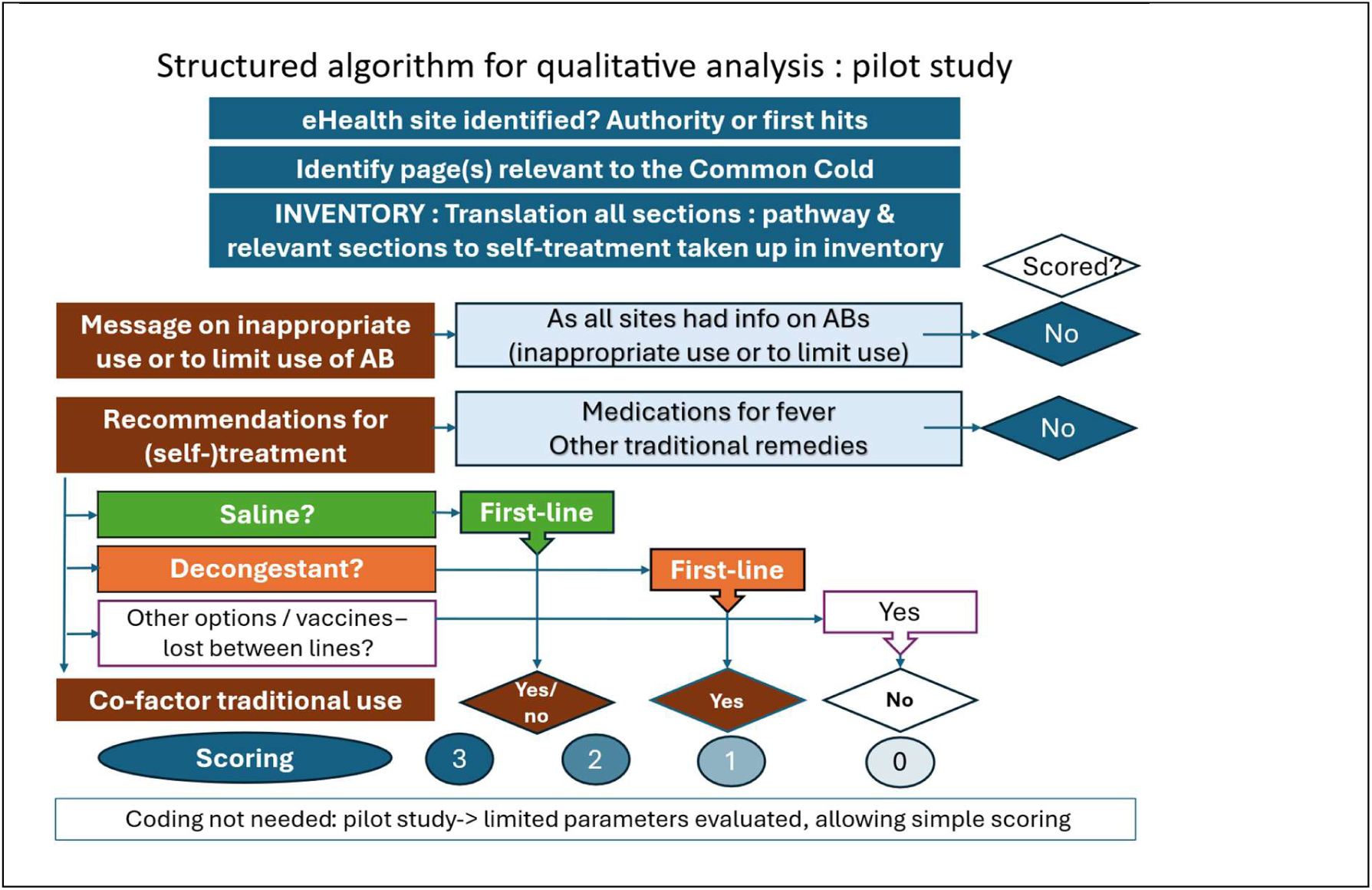
Structured algorithm followed for the qualitative analysis

Additional assumptions for the model, and their rationale, included:

- **Antibiotics**: because a first screening of all the e-sites revealed that all e-sites generally had a message discouraging or limiting the use of ABs in the common cold, this parameter was not taken up in the scoring model for the correlation assessment.

- **Traditional use of SNI:** traditional habits prevail in the ultimate choices of patients to follow, or not to follow, a eHealth recommendation [26,32]. Hence, it was assumed that ABC increases if, there is no traditional nasal saline use or clearcut persuasive SNI recommendation.

- **Non-scored treatment options:** other remedies (such as steaming or humidifying air) and medications for fever (usually short-lasting) or for pain alleviation (acetyl salicylic acid or non-steroidal anti-inflammatory drugs) were not taken up in the scoring model. There is no evidence from randomised controlled clinical trials that these treatments are effective in easing respiratory symptoms or relieving acute URTIs, and reducing ABC [36,37,38,39].

- **Vaccination:** was not systematically searched for as a co-variable, as it does not reduce the incidence of common colds and URTIs, except for influenza and COVID-19. Therefore, it was only mentioned if found on the e-sites analysed, via explicit mention or via (automatic) deviation from this target word to the “influenza” or “COVID-19” page on the e-site.

- **OTC availability of ABs** was not considered, as this information is not readily deducible from the e-Health sites or the ECDC-data. The relevance is addressed in the Discussion.

- **SNI formulations**: Given that most of the eHealth sites recommended saline nasal treatment (drops, spray, rinse, irrigation), but not saline gargling, we further referred to SNI in the text. An exception was Italy, proposing only gargling (not nasal rinse/irrigation) with saline, in a last-line position on the common cold e-page [40].

### Scoring

Scoring and grouping of the countries were performed by main recommendations and traditional use of SNI for the common cold: see **Table 2**. This approach allowed to prioritise the e-recommendations for SNI first-line, decongestants first-line ‘without’ SNI recommendation, or recommendations proposing, or focusing on, other medicines, vaccination and complications (no or no-clearcut SNI recommendation). With “no clearcut recommendation” is meant that SNI was solely mentioned as an option somewhere lost between the lines, was not proposed for relief, or only at the very end of the internet page (so unlikely to be read), or only recommended for babies, toddlers or kids (so SNI unlikely be used in the child once outgrown the toddlers’ age). SNI is unlikely to be used by the e-reader in the case that there is overwhelming or first-line information on decongestants or other medicines, vaccination and/or complications telling to seek a doctor.

**Table 2.** Scores used to categorize countries based on the recommendations on the eHealth portal for the treatment of the common cold.

| Category | Score | Features assessed |
| --- | --- | --- |
| Group 1 countries | 3 | SNI (+/- steaming) recommended first-line for common cold, irrespective of traditional use of saline for common cold |
|  | 2 | SNI traditional, but no clearcut support by the national health authority first-line, yet consistently by medical associations, health care workers, media and other communications |
| Group 2 countries | 1 | Decongestants recommended first-line<br>No saline recommendation<br>Yet traditional use of SNI for the common cold |
| Group 3 countries | 0 | Emphasis on medicines, vaccination and complications<br>No (or no clearcut) saline recommendation<br>SNI is not traditional for common cold |

Taking the traditional use of nasal saline as a co-variable into account, the approach allowed to convert the qualitative findings from the inventories into four major numerical scores and to group the countries, stratifying them by the four scores (**Figure 2)**: the higher the score, the lower the expected ABC. For the details on attrition of the scores to the countries in the main and sensitivity analysis, see Supplement, **Table S3** and **S4**.

### Statistics

Basic descriptive statistics (median and means +/- standard deviations) and correlations were calculated, using the ‘Statistics Kingdom’ calculators for the Pearson and Spearman correlation coefficients, the Mann‒Whitney U test for comparison between 2 groups, and the Kruskal‒Wallis test for continuous variables, alpha-corrected (Bonferroni) for the comparison between 3 groups.

## Results

### 1. Qualitative analysis of the authorities’ eHealth site

#### 1.1. Messages on ABs

Generally, most eHealth sites discouraged the use of ABs for use in common colds yet used different messages (**Table 3**): they explained the viral cause, and/or communicated that ABs are not of help; or are “needed for”, or are “only effective in bacterial infections”; or that ABs “need to be used properly”, so possibly evoking different perceptions with patients.

**Table 3.**
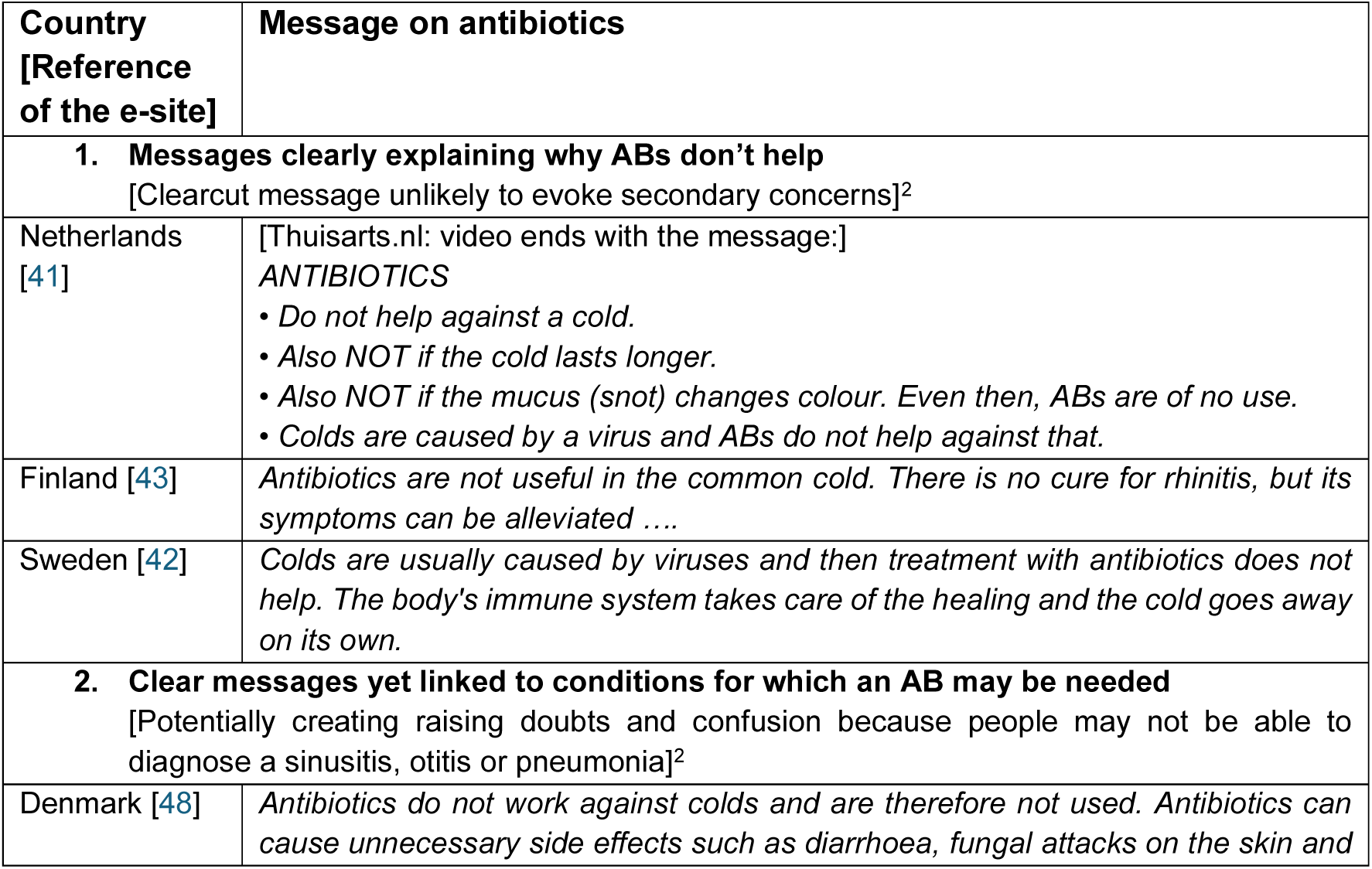
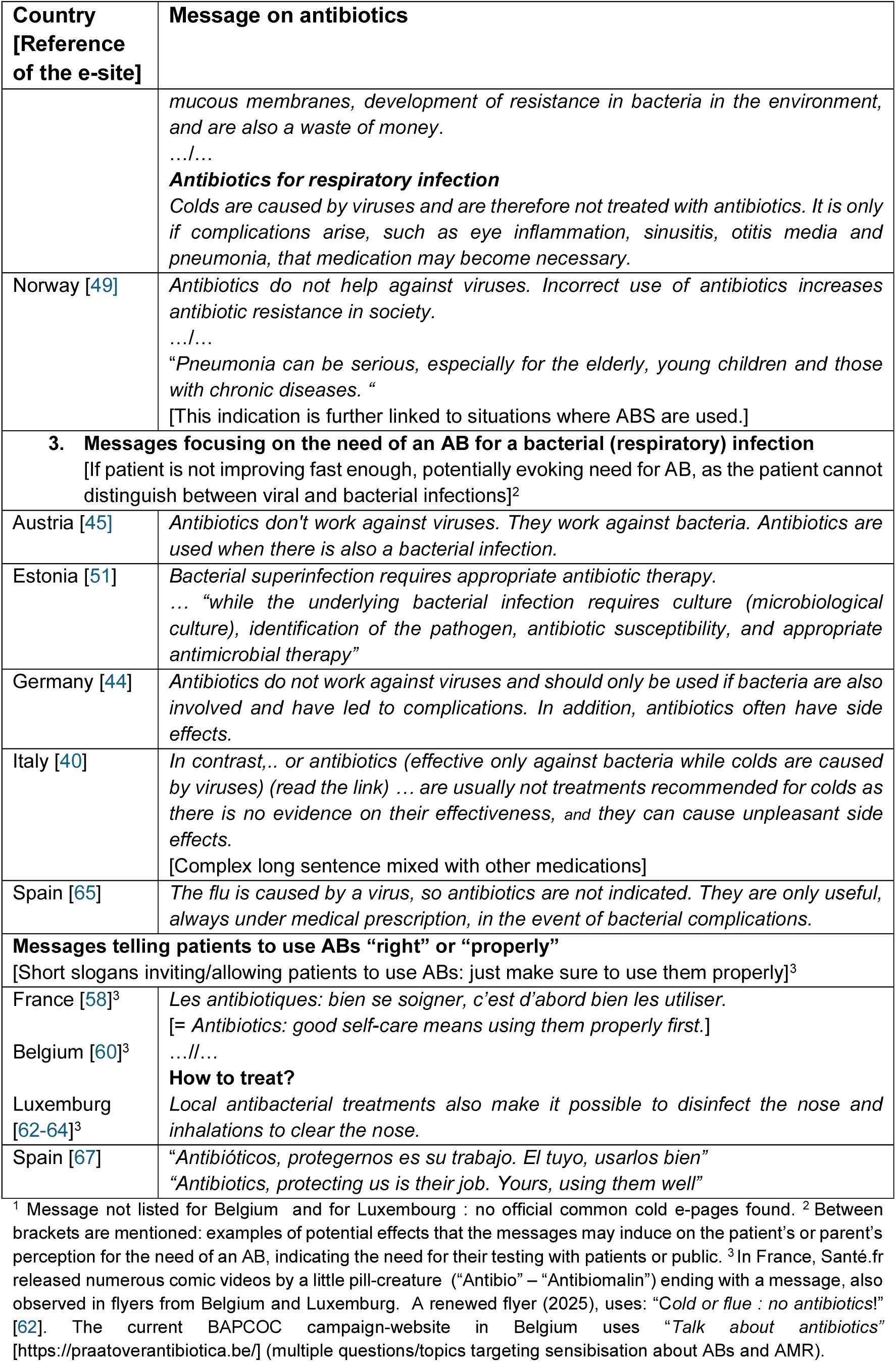
Messages on ABs identified on the common cold webpages of Public eHealth sites. [between brackets: suggestion of the potential connotations patients may give with regard to the need for an AB].^1,2^

#### 1.2. Treatment Recommendations and Subgroups by Scores

The treatment recommendations were highly different across the 13 eHealth sites. Three main groups of countries were identified according to the e-recommendations, as further reviewed and scored below (**Figure 3**). For the e-pages’ structure and translated contents, and the details of the qualitative analysis, see the inventory in **Table S2**.

**Figure 3.**
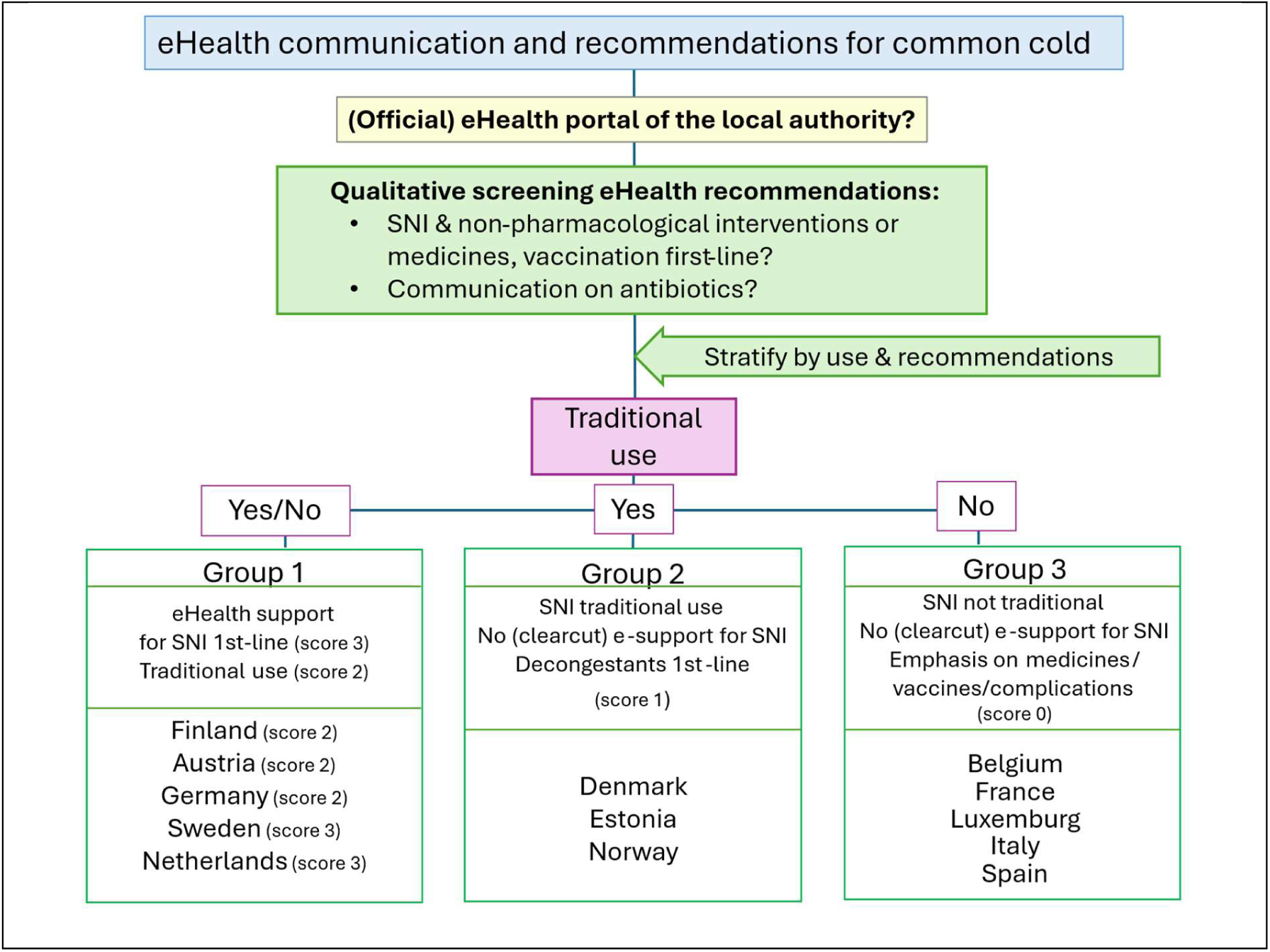
Stratification of the countries by eHealth recommendations for SNI according to qualitative screening of the eHealth site(s) (for scores see Table 2).

For the following 5 countries, e-communication was found to be in support for SNI first-line:

a. by a clear first-line positioning based on their official e-site, directly accessed upon the first hit for ‘common cold’ : the Netherlands (Thuisarts.nl [41]) and Sweden (1177.se [42]) – given score 3;
b. by persuasive recommendation of SNI for “relief” on the official e-site, after briefly discussing the limits of decongestants (Finland [43]), or the e-site directly linking the treatment options to another official e-site for treatment recommendations, recommending saline (Austria [44]). For Germany, the saline option was not mentioned on the official e-site [45] but amply identified on other e-sites by health care professionals (both private doctors and associations). These countries having historical tradition of using SNI [46,47] were given a score 2.

Despite being countries with traditional SNI use, Denmark [48] and Norway [49] received only a score 1, as clearly recommending decongestants first-line; the Norwegian page mixed the communication on saline and medicated drops, likely confusing the reader if not visiting the more specific children-oriented page where saline is proposed [50]. Estland was given a similar score because of its first hits recommending decongestants first-line (arts.ee [51]), OTC access [see Discussion], yet traditional use and clear SNI recommendations by professional organisations [52]. This country was therefore assigned a score of 1 but was augmented to a score of 1.5 in the sensitivity analysis.

All other countries (Luxembourg, Belgium, France, Italy and Spain) were assigned a score 0 because the messages of these eHealth platforms were either poor, or overly complex, with a lot of information on various medicines, vaccination and possible complications. Usually there were no clearcut eHealth guidelines for selfcare with SNI for the common cold; the message on SNI was lost between the lines (only found after repetitive searches), or the message was located at the very end of the webpage, so overflooded by other information on the webpage about, for instance, other options called effective/offering relief or complications, so creating disease mongering [31]. Italy [40] only proposed saline gargling for relief from sore throats and stuffy nose in “some”, last-line after emphasis on relief by medications, thereby also proposing ABs for bacterial infection in the same paragraph. For France, access and messages were overly complex and confusing due to difficult unstructured access, with various e-sites popping up, such as ANSM [53] and VIDAL [54] and Santé.fr [14 hits for ‘rhume’ on this site]: rather than a straight forward well-structured site on the common cold, focus was on restriction of oral decongestants [55], RSV/bronchiolitis [56]; often was saline only proposed at the very end of the page, linked to child treatment, and/or just for moistening the nose [57, 58, 59]. On the site Santé.fr, a video aiming at restricting antibiotic use, proposed by a comic pill-like creature “Antibio Malin” proposed antibiotics with the slogan “good self-care means using them properly first)” – a message seemingly confusing as it would be smart to use AB, especially when reading elsewhere under treatment that “*Local antibacterial treatments also make it possible to disinfect the nose and inhalations to clear the nose”* [58]. Belgium and Luxemburg had no persuasive e-site targeting the patients seeking information (see **Table 1**, **Table 2S** and **3S**). The same French AB message was found in Belgian flyers [60], coordinated by BAPCOC [61], and older flyers from Luxemburg (SanteSecu.lu) [62]: these recommended SNI between the lines in 2012 [63], yet not observed in later versions and 2025 [64]. In Spain (the country with highest ABC in our analysis), hits were meddled by links going to platforms from all over the world, including e.g. Mexico, Columbia, Chili, Peru, Pan American Health Organisation, and to many sites in English originating from the United States (e.g. Mayo CLINIC and CDC), popping up with their Spanish translations The official e-site Ministerio de Sanidad y Politica Social (Sanidad.gob.es) with 118 hits related to “resfriado”, did not allow to identify simple clearcut guidelines for the common cold (inappropriate use of ABs mentioned last-line)e [65], in contrast to the popular, highly visited commercial site “Sanidad.es”, not recommending saline and without a message on inappropriate use of ABs [66]. The latter message seems rather covered by a separate agency (AEMPS) [67].

### 2. ABC Use and Correlation to SNI Recommendations

#### 2.1. ABC (J01, J01C and J01F)

**Table 4** lists mean ABC (+/- standard deviation), median and *p*-values per group of countries for the global antimicrobial class J01 and its subclasses J01C and J01F (relevant to (U)RTIs). ABC J01 is illustrated in **Figure 4**. The differences in ABC J01 were statistically significant between Groups 1 and 2, in comparison with Group 3 countries (Mann-Whitney U test, *p* = .007 and *p* = .036, respectively). Group 1 countries also showed a significantly lower overall ABC than Group 2 (*p* = .036). The differences in ABC J01 were also statistically significant between the three country groups according to the Kruskal-Wallis test for continuous variables (alpha-corrected, Bonferroni correction; *p* = .017). As shown in **Table 4**, ABC was lower, and so the differences in ABC less outspoken, in the subclasses J01C and J01F (for significant differences, see **Table 4**).

**Figure 4.**
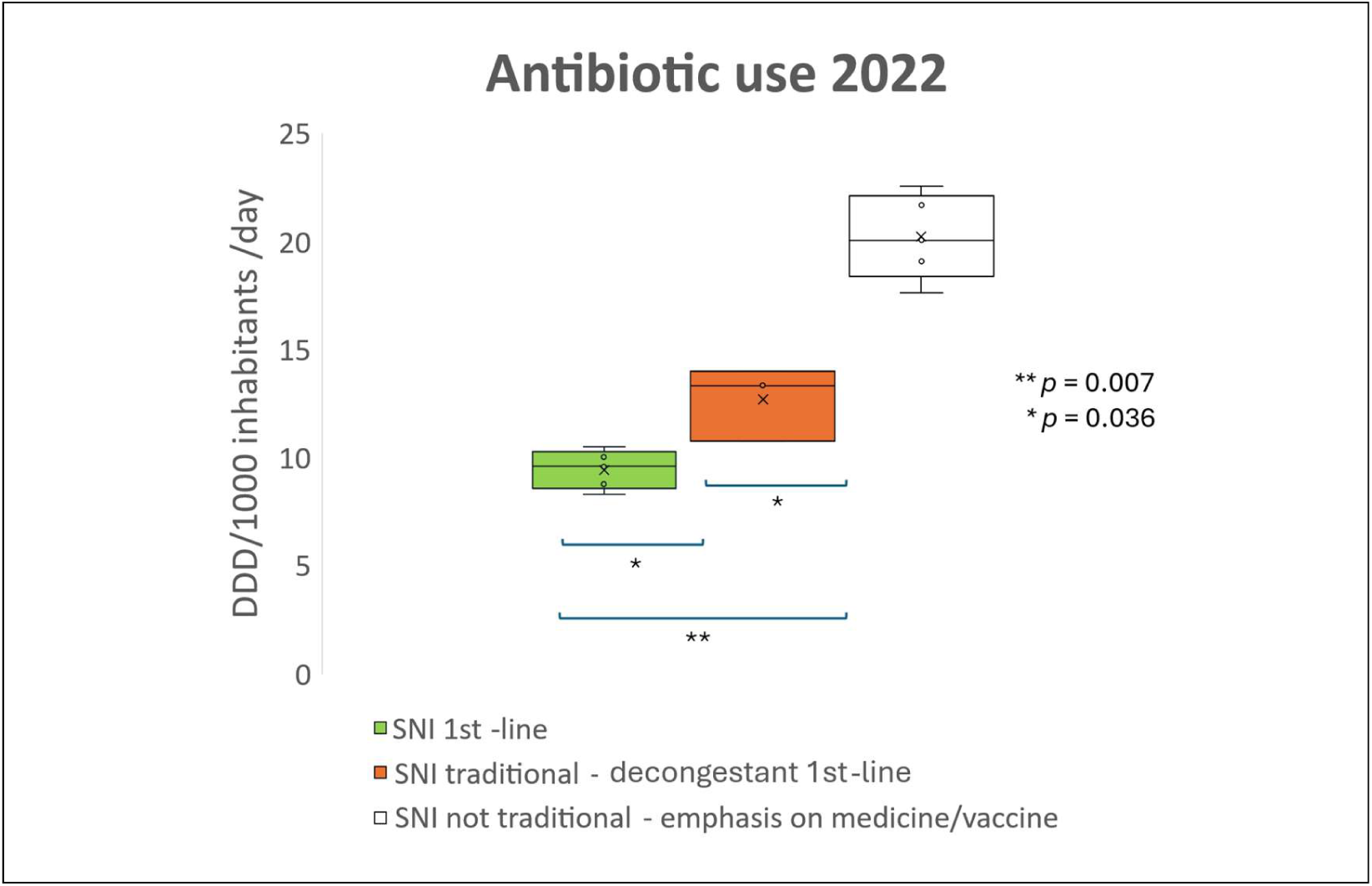
AB consumption 2022 in 13 European countries stratified by eHealth treatment recommendations (box plots).

**Table 4.**
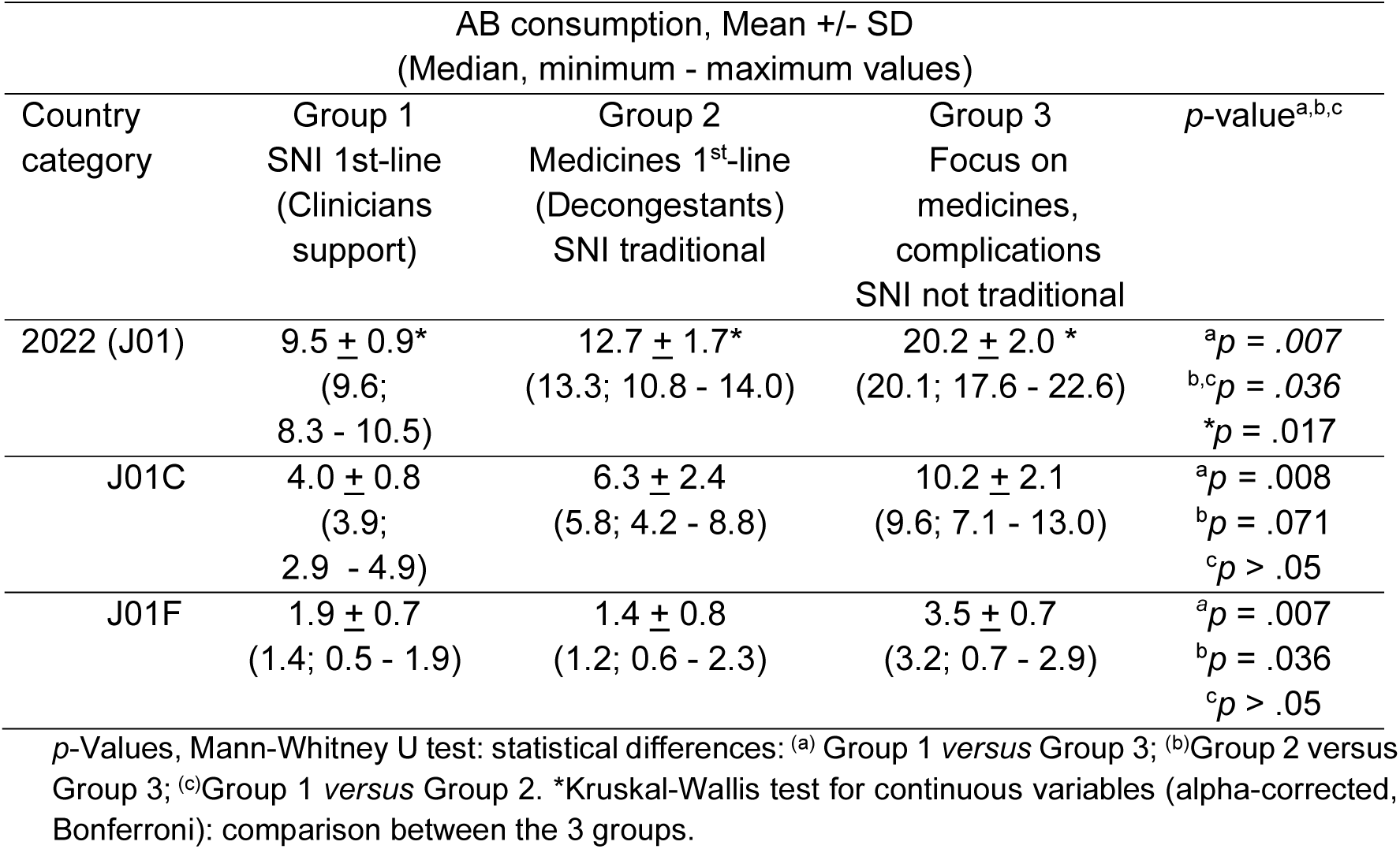
Antibiotic consumption (ABC) 2022 per group of countries stratified by recommendations/SNI traditional use for the common cold: DDD per 1,000 inhabitants per day, community sector excluding hospitals; ATC class J01 and subclasses J01C and J01F.

| Country category | AB consumption, Mean +/- SD<br>(Median, minimum - maximum values) | | | $p$-value<sup>a,b,c</sup> |
| --- | --- | --- | --- | --- |
|  | Group 1<br>SNI 1st-line<br>(Clinicians support) | Group 2<br>Medicines 1<sup>st</sup>-line<br>(Decongestants)<br>SNI traditional | Group 3<br>Focus on<br>medicines,<br>complications<br>SNI not traditional |  |
| 2022 (J01) | 9.5 ± 0.9*<br>(9.6;<br>8.3 - 10.5) | 12.7 ± 1.7*<br>(13.3; 10.8 - 14.0) | 20.2 ± 2.0 *<br>(20.1; 17.6 - 22.6) | <sup>a</sup>$p = .007$<br><sup>b,c</sup>$p = .036$<br><sup>*</sup>$p = .017$ |
| J01C | 4.0 ± 0.8<br>(3.9;<br>2.9 - 4.9) | 6.3 ± 2.4<br>(5.8; 4.2 - 8.8) | 10.2 ± 2.1<br>(9.6; 7.1 - 13.0) | <sup>a</sup>$p = .008$<br><sup>b</sup>$p = .071$<br><sup>c</sup>$p > .05$ |
| J01F | 1.9 ± 0.7<br>(1.4; 0.5 - 1.9) | 1.4 ± 0.8<br>(1.2; 0.6 - 2.3) | 3.5 ± 0.7<br>(3.2; 0.7 - 2.9) | <sup>a</sup>$p = .007$<br><sup>b</sup>$p = .036$<br><sup>c</sup>$p > .05$ |

$p$ -Values, Mann-Whitney U test: statistical differences: <sup>(a)</sup> Group 1 *versus* Group 3; <sup>(b)</sup> Group 2 *versus* Group 3; <sup>(c)</sup> Group 1 *versus* Group 2. \*Kruskal-Wallis test for continuous variables (alpha-corrected, Bonferroni): comparison between the 3 groups.

#### 2.2. Correlation of ABC with SNI Recommendations/Traditional Use

ABC strongly inversely correlated with recommendations (+/- traditional use) for SNI, when calculated by the Pearson correlation coefficient (*r* [Confidence Intervals] = ^_^0,905 [CI: 0.972; -0.706], *p* < .001)) and even more so by Spearman correlation (*r*_s_ = -.945 [CI: -.989; -.738]; *p* < .001). The inverse correlation remained high for the AB subclasses of interest to (U)RTI (J01C *r_s_* = -0.845, *p* < .001; J01F: *r_s_* = -0.799, *p* < .001, respectively). In a first sensitivity analysis [**Table S4-A**], downscaling the maximum scores by no longer differentiating between scores 2 and 3, so simplifying to 3 categories of countries (scores 0 – 1 – 2), the strong inverse correlation remained retained. Vice versa, after fine-tuning the scores with +/-0.5 extra points, the correlation remained significant (*r_s_* = -.927 [CI: -.978, -.768]; *p* < .001).

## Discussion

In line with the global call for prudent use of ABs [1–3], most of the common cold pages of 13 public European eHealth sites carried a message discouraging the use of ABs. Yet, these messages were highly different, so are likely to evoke different patients’ perceptions and needs, calling for more in-depth investigation. For instance, the proposition that ABs are needed for bacterial infections may create feelings of uncertainty, as patients and parents easily experience confusion about which symptoms need treatment by ABs, even more so if the physician is unclear in his/her communication [68,69].

Using, however, a simple scoring system and country stratification by prioritisation of the recommendations for the common cold, taking also traditional nasal saline use into account, a surprisingly powerful inverse correlation was identified between SNI recommendation and ABC in the community. This suggests that treatment recommendations, with first-line recommendation of SNI as affordable and effective solution for the common cold, may help to change the patient’s behaviour towards ABs, so help reduce ABC. The results are in line with the recent findings that a message of fear against the use of ABs, or a GP’s requests to delay an AB, would not work well with the public unless if combined with reassuring information offering effective symptom control [68,70,71,72]. A problem of the national or European programmes focusing on ‘education’ of public, patients and physicians about (in)correct use and adverse effects of ABs [e.g. 73,74,75] may thus be they leave patients without an effective solution for their complaints and their perceived needs [28], hence sustaining barriers, as further discussed. This is also suggested by other observations: in Italy, 74 % of Italians participating in a recent survey [76] knew that ABs were an ineffective treatment against the common cold, yet they still relied on numerous medicines and self-medication with ABs – particularly in the south of Italy [77,78,79]. A recent study of educational messages on AMR disseminated in social media revealed that these had raised high awareness about AMR but failed to change AB practices [80]. A study across Europe 2014-2018 found statistically significant decreases in total ABC in only eight ESAC-Net countries [81]: six of these were rated in our analysis as recommending SNI or using traditionally SNI for the common cold, further supporting that offering SNI persuasively as a solution may function as a facilitator to change the patients’ behaviour to AB use.

Although a role for public internet education on ABC was identified more than 2 decades ago [82], the impact of well-accessible eHealth on today’s patients’ health advice seeking behaviour should not be underestimated. In the Netherlands, the e-site Thuisarts.nl reached 80 million visitors in 2022 [83], while it was already shown in 2016 that – after launching an official eHealth site by the Dutch authorities, the healthcare usage decreased by 12% after 2 years [84]: the findings showed that trustful eHealth information to patients can be effective in improving self-management and reduced healthcare usage in times of increasing healthcare costs. In this study, it was observed that the countries with well-structured and easily accessible e-site information on the common cold page (Austria, Finland, Germany, Netherlands, Sweden) generally had a low ABC in our study. This is in line with a former study across Europe finding that the citizens’ subjective perception on AB use correlated with the countries’ ICT development index (ICT access, use, and skills) (*r* = 0.82), thereby achieving a moderate negative correlation with ABC (*r* = -0.41) [85,86]. As implementation of AB policies was however found to be without significant effect, the study acknowledged that additional factors play a role [86].

In fact, several barriers against reducing ABC have been identified [87], that can be addressed by eHealth communication by providing treatment recommendations, overlooked so far:

(1) Difficulties with accepting to manage self-limiting infections with simple rest and symptomatic ) treatment”: this common patient attitude implies that eHealth sites should propose convincingly accessible selfcare that works, as to engage the patients to follow the proposed e-recommendations: inappropriate self-mediation can lead to undue consultations and therapy [88]. Fear, anxiety, negative emotions and uncertainty engage patients in self-medication with ABs [26,89].

Proposing paracetamol and ibuprofen [70] may not work to reduce ABC, as shown in an earlier clinical study [24]. Many e-sites offered this option, yet regarding the countries with lowest ABC 2022 [8], the Dutch e-page didn’t mention this option [41], while Sweden reassured that treating fever is not always needed, before proposing an antipyretic [42]. Vice versa, the latter countries proposed saline, often overlooked as valuable first-line option for treating common colds on other e-sites. Barriers against the use of nasal saline use include the beliefs that it is ineffective, unnecessary, dangerous or problematic to use [90,91]. Moreover, recent reviews of treatments for the common cold [92,93], surveys or reviews on parents’ beliefs and pharmacist-proposed remedies [26,28,94,95], or studies assessing selfcare in Europe [96,97] failed to address the role of SNI, or limited its use to children [92,93,98]. Rather the way around, there is a tendency to discredit or minimise the role of nonpharmacological options by doctors, saline inclusive, by stating that there is no or insufficient experimental evidence for them [100,46,99]. It is however to note that the Netherlands already integrated SNI in its national clinical standard guidelines for primary care physicians in 2014 [100]; this recommendation was consistently implemented on many other professional sites (see Inventory in **Table S2**); in the Scandinavian countries, SNI is a common traditional practice, with high proportions of healthcare professionals recommending it for their patients [47]. Today, both from efficacy and safety perspectives, SNI can be safely recommended for selfcare in URTIs, its symptoms overlapping with COVID-19 [24,101,102]. Further, compared to counselling alone on ABs (found to reduce ABC), nasal saline resulted in more clinical benefits [24], making it a valuable candidate for persuasive recommendation on eHealth sites.

(2) The difficulty to translate “theoretical awareness in actual prudent prescribing behaviour”: this is attributed to the “perceived clinical risks, the relationship with the patient and the perceived patient demand for ABs”; it leads to the idea “it’s better to prescribe/take too much than too few”. Patient expectations and diagnostic uncertainty are unlikely to be solved by the eHealth messages on ABs (Table 1). They have been shown to exert significant prescribing pressure on physicians [26–28,29,103,104,105]. The ratios of AB prescribing in response to a patient’s request for ABs, assessed across 18 European countries, were found to be enhanced, yet highly variable across countries [29]. Applying them in our model (ABC data from ECDC 2022) resulted in a noticeable correlation (*rs =* .384, *p* = .195), yet not as strong as found for the eHealth treatment recommendations in this study.

In this regard, restrictive eHealth communications on treatments, with the objective to limit their use, may also need reconsideration: while generally being utile [106], if inappropriately formulated or positioned, such messages may possibly - unwillingly – create uncertainty with the patient, and disease mongering [31,107]: instead of guiding the patients to more restricted medicine use, the primary messages that say “it helps, but that it should be used properly “(as is the case for ABs on many eHealth sites) or “don’t use it too long” (such as with decongestants) may rather drive patients to seek the medicine: such messages may create the perception that the medicine is needed, or does better for getting relief. In fact, it is like with children: continuously pointing the finger and forbidding things, can be counterproductive, especially if no effective alternatives are offered [108,109].

Our analysis revealed that the first-line proposition of decongestants by the authorities of Norway and Denmark was parallelled by an enhanced ABC versus other Scandinavian countries recommending saline (Sweden and Finland). In line with the Norwegian e-recommendations [49], currently 700,000 Norwegians would be dependent on nasal decongestant sprays (population of 5.5 million, so 1 in 8) [110]. The mechanism for increasing ABC may not just be through creating false expectations and the subsequent need to seek the doctor’s advice, if there is no improvement beyond the allowed duration of use. Other causative factors may be rebound congestion [111], and (from pathophysiological perspective) the inhibition of the cilia in the nasal epithelium, so inhibiting clearance of the infectious pathogens [112].

(3) “Lack of confidence in existing guidelines”: the selfcare measures need to be persuasively proposed to create confidence, such as found in our analysis by ‘Thuisarts.nl’ [41] or “1177.sv” [42]. This is the more important for countries with high ABC due to self-medication with ABs, because for selfcare, people rely rather on their habits than the internet [32]. In Italy, a country with high ABC and self-medication with ABs [77], the current e-message by Instituto Superiore di Sanita only offers SNI as a last option, saying “some” (“alcune persone”) may benefit from “gargling salt water, this after firstly proposing medicines, decongestants inclusive, that “work” for the common cold [40]. Such message is unlikely to convince AB self-medicators to selfcare URTIs with SNI. The e-recommendation need also to apply in general: if just recommended for children, adult patients are unlikely to follow the recommendation, while doctors in many countries tend no longer to recommend SNI, or parents may be less convinced to continue SNI, once the toddler age is past. As shown by independent research, parents and teachers often believe a child can go back to school whilst feeling unwell, if taking an AB [113]. Persuasive recommendation of saline by clinicians beyond toddler’s age is needed, in view of the facilitators for, and barriers against nasal saline [90,91], and the strong decrease in use observed by the age of children (to 15% or less in over 1 years of age) [91]; significantly more parents understood the importance of NSI when their pediatricians considered NSI effective [91].

Overall, these barriers suggest that providing only fear or restrictive public eHealth messages may possibly be of little help to convince patients for changing their habits, perceptions, and the needs for ABs, unless the eHealth communication, as well as clinicians’ information to parents and patients, also delivers a clearcut persuasive message offering an accessible, effective solution such as SNI. Just talking about ABs, raising awareness about AMR, may not suffice. In fact, this should not be surprising, because (promoting) restrictions without offering a clearcut solution are well-known to be counterproductive in education [108,109], disease mongering [31], research on communication [25] and other areas of healthcare [114]. The findings call for study of eHealth messages on ABs, saline and other treatments in the national recommendations and One Health programmes. In these regards, **Table S5** summarizes several aspects identified from this study. They include, for instance, the impact of proposing benefits of nasal saline, recently identified as facilitators to change patients’ perceptions [90] and the impact of readily providing the instructions how to make saline and/or how to use it, on the e-site. Such instructions were found on the Dutch, Finnish and Swedish official e-sites [41,42,43], all countries with low ABC. But also the study of the patients’ attitudes to recommendations versus empowerment of the AB message is warranted: for instance, to overcome barriers against its use (90), messages may be tested highlighting the benefits/efficacy of SNI (clinical and reduction in ABC, both adults and children), ease of preparation, availability in the pharmacy, and safety for selfcare, even if overlapping with COVID-19 [19,21–24,101,102], this versus the current communication often limiting the role of saline to simple cleansing of a stuffy nose in children. Also, the effects of the explicit proposition of medicines that “work well for relief”, yet are restricted in use, such as the decongestants, need evaluation: what are the patients’ expectations for an AB prescription, if their respiratory symptoms are not resolved within the allowed days of use?

## Limitations of the Study

We based our findings on what is directly found on the internet in a way that consumers would find their information on the internet, when seeking a solution for the common cold on eHealth sites. The information reviewed by us may not necessarily reflect the official clinical recommendations to the physicians. Moreover, the numerical scores used for the conversion of the qualitative information from the eHealth site to calculate the correlation were based on assumptions in which SNI was given a central role, while possibly also other (non-assessed) remedies and recommendations in other diagnoses may contribute to the ABC. Yet, the findings were surprisingly highly significant, and the correlation was retained for the subclasses of ABs that are mostly relevant to URTIs, albeit at lower significance levels. These correlations were also maintained when using the recently released European ABC data 2023 [115] (***r_s_*** = -0.9422; ***r_s_*** = -0.9862 in the main and finetuned-sensitivity analysis, respectively; **Table S3-B**). To note, while trends in ABC since 2001 were calculated [115], we found hardly or no rebound versus 2019 in countries recommending SNI (Sweden, Austria, Germany, Finland and Netherlands), while ABC further increased in Estonia, Denmark and Norway, corroborating the findings of this study.

We did not specifically address, nor score, the specific recommendations for the common cold in children. These appeared to be well addressed, with recommendation of SNI, via the e-sites in the Netherlands [41], Sweden [42], Finland [43] and Austria [44], while not in Germany [45] and Denmark [48], and with no clearcut recommendations readily found in the countries with high ABC (see also Inventory in **Table 2S**). In Norway, the separate topic “Congested nose in children” proposed saltwater drops in the same section as decongestants ending with a confounding note for parents: “It is not certainly documented that these medicines have an effect on children” [50]. Based on national prescriptions registers, Scandinavian countries have observed a decline in ABC in children between 2006 and 2017, particularly during the first two years of life; the lowest use was observed in Norway yet showing the lowest decline in ABC [116]. A recent assessment implicates fragmented markets for older ABs and child formulations in Scandinavia, whereby countries with similar resistance patterns and prescribing cultures (e.g. Norway and Sweden) prioritize different formulations and dosage strengths in children [117]. So, apart from the study of eHealth messages how best overcoming parents’ barriers against using SNI [90,91] and misbeliefs about ABs [113], also other aspects need consideration.

Many other factors related to, or other than evaluated in this pilot study, may play a role. As proposed for further study in **Table S5**, these include a.o. the accessibility of the e-site, the exact message on ABs, the messages on when seeking the doctor’s advice on the official e-sites, but also the information by commercial promotion on often more appealing pseudo-official (pharma-driven), para-pharmaceutical and pharmacy e-sites, and the consistency of the recommendations throughout local and European professional clinical guidelines, as well as across related diagnoses (URTI, sore throat/ acute laryngitis, ear pain/otitis, acute bronchitis and acute (rhino)sinusitis): these diagnoses have also been shown to be subjected to highly different rates in AB prescribing [13]. As these ailments often start with cold symptoms, while share benefits with SNI [24,101,118,119,120], there may be a spill-over effect of persuasive first-line SNI recommendation for the common cold, leading to delayed or lower ABC also in these indications. SNI was recently found to be an acceptable alternative to ABs for patients with sinusitis, when being proposed by their primary care physician: providing instructions on how to prepare and administer were relevant [121,122]. Moreover, similarly, as for URTIs [38,39], patients experienced steam as being rather ineffective (only relief the headache) [121], while more benefit was obtained with SNI [123].

Other aspects may include physician, pharmacist and student education: positive studies usually relate to indications generating less ABC (urinary tract, dental and eye infections) [124]. A recent systematic review raises the question as to the usefulness of clinician education on AB use, as doctors are generally aware of the predominant viral origin of URTI [10]. Recent surveys among medical students across Europe [125,126] revealed uncertainties and a need for further education on ABs, regarding the recognition of clinical signs, severity and markers of infections that need an AB, and the differentiation between bacterial and viral URTIs - typical uncertainties shared by patients, parents, pharmacists and doctors alike.

It can also be criticised that the countries with a higher ABC, selected in our pilot study, may have been influenced by higher self-medication with ABs. Countries with high levels of regulations observe less ABC [127]. More precisely, self-medication practice with ABs is common in countries where ABs are available OTC in the pharmacies due to a lack of enforcement or penalisation of such practices by the national regulators. This may have been the case for Spain and Italy [79,128,129,130], but not for Belgium and Luxembourg [87]; it is unlikely the case for France, which implemented and enforced AB stewardship activities well before COVID-19: these led to a substantially declined ABC, yet remaining 30% higher than the European average, and 2-3-fold higher than in Scandinavian countries [131]. Yet, according to our analysis, France also recommends nasal antiseptic sprays, or antibacterial treatments, as effective options for the common cold, so possibly reinforcing the perception for needing an AB. The case of Estonia, another country with easy OTC access to ABs, proves that the OTC-availability of ABs is not necessarily the most determining barrier against changing the patients’ habits: non-prescription use of ABs was found to be much less common than in Spain and Greece [79], while the 2022 ECDC-data used in our study [8] revealed that the ABC is also lower than in Belgium and Luxembourg, two countries that fully lack OTC access to ABs. The latter two countries, however, also lack traditional use and/or persuasive recommendations on nasal saline in contrast to Estonia [52]. In fact, a former factor impacting on ABC, called Hofstede’s ‘cultural’ values, [132] is both consistent with the effects of OTC-AB accessibility and SNI recommendations, as used in the model: they integrate indulgence versus restraint dimensions, uncertainty avoidance, pragmatism and orientation towards future rewards, all parameters expected to be influenceable by eHealth communication and (traditional) use of SNI. The findings corroborate that the persuasive recommendation of SNI for the common cold may act as a facilitator for ABC reduction, irrespectively of OTC access to ABs.

Finally, factors beyond the scope of this study include: (a) outside Europe, the rising income and increased access to ABs in low- and middle-income countries [133]; (b) the AMR itself, driven by AB (over)use in farming [134] and hospital use [e.g. 135]. Hospital use across Europe is much lower than in the community [115] yet did not correlate with our model’s public eHealth recommendation scores (**Table 3S-C**). This is not surprising, as hospital ABC follows complex metrics across the member states [136]: it is influenced by national and local hospital guidelines, the risk of nosocomial/hospital-acquired infections, and the physicians’ prescribing practices: as in the community, similar uncertainties, such as the ‘lack of microbiological results’ and ’no access to AB susceptibility patterns’ remain major barriers to more restrictive AB use [137,138]. However, in the hospital, the fear of missing bacterial infections or sepsis prevails, as does the need to prevent complications, particularly in the co-morbid patient. So, dynamics differ when compared with the ABC in the community.

## Conclusions

This pilot study identified highly different treatment recommendations for the common cold on public eHealth internet sites of 13 European countries, with a strong inverse correlation between their prioritization of SNI recommendation on the e-sites and the ABC in the community: the lowest ABC was found in countries proposing SNI as a first-line option; the highest in absence of such recommendation and/or SNI traditional use, yet upon emphasis on decongestants, other medicines or complications. The findings suggest that the perceptions and needs of the many patients seeking advice on the internet should be better met by offering an effective solution. SNI is a worldwide-affordable intervention and a traditional hygiene practice with proven benefits and without significant risks. Albeit many aspects affect ABC, the results call for more in-depth study on the role of recommendations of remedies, such as SNI, in the common cold and URTIs, and of the public e-health communications in general, in the action plans to educate patients, combat AB overuse, and to help achieving Sustainable Development Goals.

## Data Availability & Ethics Statements

**T**he ECDC Eurosurveillance data on antibiotics (community sector) were used as published by Ventura-Gabarró in the Eurosurveillance Journal November 2023. All data used has been provided in the supplement to this publication. No other data was used. For the qualitative assessments of the recommendations, all links consulted have been inventoried per country in the Supplement. The work also adheres to the international standards for authors as proposed by the Committee on Publication Ethics (COPE).

The following abbreviations are used in this manuscript:

## Abbreviations

AB: antibiotic
ABC: Antibiotic consumption
AMR: Antimicrobial resistance
ECDC: European Centre for Disease Prevention and Control
DDD: Defined Daily Doses
PSAU: Subjective Perception of Safe Antibiotic Use
ISS: Instituto Superiore di Sanita
SNI: Nasal saline irrigation (nasal saline drops or irrigation and gargling)
WHO: World Health Organisation

## Acknowledgement

This work was facilitated by a small informal network of Belgian pharmacists (Saline Solution COVID Collective), evaluating the rationale for the use of oronasal hygiene with saline nasal irrigation and gargling in COVID-19, 2020-2022 (available on request). A next step was to enhance the understanding of recommendations of oronasal hygiene with nasal saline and gargling for the common cold and upper respiratory infections (URTIs) throughout Europe. The authors thank other pharmacists and clinicians for the exchange of ideas and expertise that helped to complete this manuscript.

