## Supplemental Figure S1 Tables S1 to S4 for "eHealth Treatment Recommendations for the Common Cold across 13 European Countries: Correlation with Antibiotic Use"

**Supplement:**

**Figure 1 S. Theoretical Framework.**

Pilot study. Overview of prior theoretical framework, considered for qualitative research, categorising countries by the eHealth recommendations (SNI, decongestants , other), scoring, evaluation and correlation with ABC.

### Theoretical framework

| Phase | Key activity | Viable info? |
| --- | --- | --- |
| Design | Concept: first-line SNI recommendation affects ABC | Yes, PubMed literature |
| | eHealth site identification | Yes, Internet -> Search strategy (Table 1) + Inventory |
| | Variables: Treatment options recommended for common cold as prioritised by Authority (or relevant alternative if unavailable) | Yes, PubMed literature & pharmacy: decongestants, other nasal sprays, antivirals (vaccination) |
| | Co-variable: traditional use SNI | Yes, rationale & info from PubMed; contacts/experience |
| Pilot study | Selection of EU countries with relevant info on ABC & eHealth site | Yes, ECDC, community sector: n=13 (see Table 1S) |
| | Identify page(s) on Common Cold | Yes, translation -> Inventory (Table 2S) |
| | Stratify countries by co-variable + eHealth recommendations for 3 identified options | Yes, reproducible prioritisation (Table 2S, left column), Scoring (Tables 2, 3S,Fig,1) |
| | Evaluation ECDC values :<br>• Mean ABC per scored group of countries<br>• Correlation coefficient with scores | Yes<br>[Excell statistics]<br>+ Sensitivity analyses (Table 4S) |

**Table S1. Country Selection & Data Used:** community consumption of antibacterials for systemic use (ATC group J01) expressed as DDD per 1,000 inhabitants per day, 29 EU/EEA countries, 2019–2022 in Ventura-Gabarró · 2023 https://www.eurosurveillance.org/content/10.2807/1560-7917.ES.2023.28.46.2300604

**Countries: 13 countries**

| | Antibacterials for systemic use (ATC group J01) | | | |
|---|---|---|---|---|
| | 2019 | 2020 | 2021 | 2022 |
| Austria | 9.77 | 7.13 | 7.20 | 8.80 |
| Belgium | 19.78 | 15.25 | 16.00 | 19.01 |
| Bulgaria | 19.06 | 20.74 | 22.37 | 24.19 |
| Croatia | 16.93 | 14.05 | 16.22 | 18.18 |
| Cyprus | | | | |
| Czechia | | | 11.49 | 13.93 |
| Denmark | 13.44 | 12.51 | 12.61 | 13.33 |
| Estonia | 10.24 | 8.81 | 8.66 | 10.78 |
| Finland | 12.56 | 9.95 | 9.45 | 10.50 |
| France | 23.34 | 18.70 | 19.86 | 22.56 |
| Germany | 11.37 | 8.95 | 8.13 | 10.03 |
| Greece | 32.41 | 26.37 | 21.78 | 31.16 |
| Hungary | 13.27 | 10.03 | 10.82 | 13.35 |
| Iceland | 17.96 | 15.38 | 15.73 | 17.47 |
| Ireland | 21.02 | 17.10 | 16.32 | 21.47 |
| Italy | 19.80 | 16.50 | 15.99 | 20.05 |
| Latvia | 12.01 | 9.97 | 10.16 | 13.45 |
| Lithuania | 14.00 | 11.89 | 12.13 | 16.17 |
| Luxembourg[a] | 19.75 | 14.84 | 14.61 | 17.64 |
| Malta | 18.71 | 14.38 | 14.11 | 21.70 |
| Netherlands | 8.68 | 7.77 | 7.63 | 8.32 |
| Norway | 13.61 | 12.76 | 12.84 | 14.02 |
| Poland | 22.23 | 17.15 | 18.84 | 22.28 |
| Portugal | 17.90 | 13.71 | 13.74 | 17.07 |
| Romania | 24.04 | 23.75 | 24.28 | 26.19 |
| Slovakia | 17.97 | 13.16 | 14.53 | 19.66 |
| Slovenia | 11.49 | 8.85 | 8.75 | 10.96 |
| Spain | 23.27 | 18.19 | 18.48 | 21.70 |
| Sweden | 10.33 | 8.92 | 8.66 | 9.60 |
| **EU/EEA[b]** | **18.53** | **15.11** | **15.23** | **18.10** |

**Not inluded: UK (no ECDC reporting)**

Austria: included
Belgium: included
Bulgaria: no developed internet instructions found
Croatia: no developed internet instructions found
Cyprus: no ECDC reporting; touristic island
Czechia: no developed internet instructions found a no local contacts to check traditional use
Denmark: included
Estonia: included
Finland : included
France: included
Germany: included
Greece: not included due to difficulty 'Greek' language; as AB can be bought OTC and use is very high, this country would only have further enhanced the correlation.
Hungary: o developed internet instructions found a no local contacts to check traditional use
Iceland : island & difficult language; no local contacts to check traditional use
Ireland: island & difficult language; no local contacts to check traditional use
Italy : included
Latvia: no developed internet instructions found combined with difficult language; no contacts to check traditional use
Lithuania: no developed internet instructions found combined with difficult language; no contacts to check traditional use
Luxembourg: included
Malta: touristic island
Netherlands: included
Norway: included
Poland: atypical with regard to AB use:" Poland has one of the highest rates of antibiotic use in Europe, ranking fifth in consumption among 23 countries in 2016. This was largely driven by over-the-counter sales of furazidine, an antibiotic for uncomplicated urinary tract infections, which accounts for 15% of systemic antibiotic consumption and is heavily advertised The increased use of furazidine, often consumed without medical indication, correlates with a 32.8% resistance rate of *E. coli* to nitrofurantoin." See [1]
Portugal: site not identified, always Brazilian sites popping up; characterized by high ABC, which is driven by OTC use – so, unlikely to change outcome
Romania & Slovakia: bo developed internet instructions found
Spain: included
Sweden: included

[1] Fal AM, Stelzmüller I, Kardos P, Klimek L, Kuchar E, Gessner A. Antibiotics Usage and Avoidance in Germany and Poland: Attitudes and Knowledge of Patients, Physicians, and Pharmacists. Antibiotics (Basel). 2024 Dec 6;13(12):1188. doi: 10.3390/antibiotics13121188.

**Table S2**. **Inventory of the Content of the eHealth sites Retrieved and Main Points of Evaluation.**

| ***Note to table ref. searches and Inventory*** |
|---|
| The primary target word for the search was “common cold”, followed by the internet extension. If there were no relevant hits, searches were subsequently performed on the target word in combination with “saline” (eventually “treatment”) and “health authority”. Texts from hits obtained at random by opening the links were translated into English by the Google Translator. If the information originated from a health authority, the first relevant hit(s) was (were) listed in Table 1. An inventory of the translated text, relevant and related to common cold from the official eHealth site or another relevant health site - regardless of whether it popped up first - was made per country; see Table S1 in Supplement 1 for the translated sections, relevant to treatment recommendations for common cold, including the nonpharmacological treatments, such as SNI(G), antibiotics (ABs) and other treatments of common colds. If needed for interpretation, additional information was retrieved from (1) representative eHealth sites supported by physicians or medical professional organisations; (2) scientific publications: if needed, PubMed was screened for “common cold” and “treatment” or “self-care” in the case of Austria [1] and Finland [2]; (3) other communications related to SNI or G on the internet, e.g., hosted by internet pharmacies, TV and lay press; the latter sources are not listed, as they were just checked for interpretation: links were opened, text searched for the target words SNI or G, and a quick translation was performed, if needed. If needed, international contacts also allowed us to verify a country’s status, and so to rate a country as traditional or not. |

| **Country +** sequence elements from algorithm | **Relevant retrieved sections & Qualitative Research Summary**<br>- Structure of the e-site<br>- Section Treatment, self-care: content translated into English<br>Other links to supportive e-info, if applicable | **Comments on Internet Search**<br>- Sequence of sites popping up<br>- Major (official) eHealth site<br>- Links to other supportive sources |
|---|---|---|
| Austria<br><br>↓<br>↓<br>↓<br>↓<br>↓<br>↓<br>↓<br>↓<br>↓<br>↓ | • Erkältung (grippaler Infekt)<br>https://www.gesundheit.gv.at/krankheiten/atemwege/grippe/grippaler-infekt-diagnose-therapie.html<br>Content of page:<br>• Erkältung: Was ist das?<br>• Wie werden Erkältungsviren übertragen?<br>• Video: Wie kann man sich mit einem Atemwegsinfekt anstecken?<br>• Wie kann man einer Erkältung vorbeugen?<br>• Welche Symptome können bei einer Erkältung auftreten?<br>• Wie verläuft eine Erkältung?<br>• Wie wird die Diagnose einer Erkältung gestellt?<br>• **Wie erfolgt die Behandlung einer Erkältung?** | Gesundheit.gv.at<br>Öffentliches Gesundheitsportal Österreichs<br><br>So this concise page does no longer recommend saline, but the focus is on the classic COVID-19 preventive hygiene measures, and some focus on medicines (paracetamol oder Ibuprofen fever), while it explains why antibiotics don’t work and that these are only to be used if there is additional bacterial infection. The page refers then under self-treatment for further information on treatment to the national health insurance site(Website der Sozialversicherung), which however states saline first-line before any other treatments. |
| Treatment | Since there are so many cold viruses, it is difficult for experts to develop targeted medications against them. Therefore, there are currently no medications that specifically work against cold viruses. However, there are various medications available to relieve cold symptoms. These include painkillers with the active ingredients paracetamol or ibuprofen. Important: Aspirin© must not be used in children under the age of twelve. It is a possible trigger for the serious so-called Reye syndrome. | As for Germany, there no longer the option for (inhalation with) saline for common cold on the Health Authorities e-Health site. Yet, it is discussed under house remedies for bronchitis. And asks the people consulting the site of the health insurance, so circumventing the evidence-based pressure of clinical experts, preferring to medicate. |
| AB message: yes | “Antibiotika wirken nicht gegen Viren. Sie wirken gegen Bakterien. Antibiotika kommen zum Einsatz, wenn zusätzlich eine Infektion mit Bakterien vorliegt.” | Yet when combining with saline, it is still recommended in German on many sites by doctor’s associations (“inhalieren mit Kochsalz”), Pneumologists, Ear-nose-throat specialists, hospitals, health insurances, pharmacies, as it is and used to be a very traditional nasal care (see Germany). Moreover a study (Gerlitz 2022) indicates that at the doctors level: only 27.4% receive antibiotics, 28.4% inhalation therapy with sodium chloride, eventually with additives, while 84.7% use home remedies before seeing the doctors. |
| Self-medication<br>↓<br>↓<br>↓<br>↓<br><br>↓<br>↓ | **What can I do if I have a cold?**<br>You can take the following supportive measures even if you have a cold:<br>• Stay in bed if you are tired or have a fever.<br>• In general, allow yourself to rest when necessary.<br>• Pay attention to fluid intake, especially in children with a fever.<br>• Don't smoke: Smoking can worsen symptoms and delay recovery.<br>• Further information can also be found on the Social Security website → link:<br>https://www.sozialversicherung.at/cdscontent/?contentid=10007.862784&portal=svportal | |

| **Country +** sequence elements from algorithm | **Relevant retrieved sections & Qualitative Research Summary**<br>- Structure of the e-site<br>- Section Treatment, self-care: content translated into English<br>Other links to supportive e-info, if applicable | **Comments on Internet Search**<br>- Sequence of sites popping up<br>- Major (official) eHealth site<br>- Links to other supportive sources |
|---|---|---|
| ↓<br>↓<br>Saline first-line<br><br>Other support for saline? Yes | **Here's how you can help alleviate your symptoms:**<br>• Take it easy and stay in bed if you feel weak or have a fever.<br>• **Inhaling with steam, nasal douches with salt water and gargling with sage tea are often found to be helpful**<br>• Do not smoke. Smoking worsens the symptoms and delays healing.<br><br>SNI/inhalation is supported e.g. in<br>https://www.netdoktor.at/krankheiten/erkaeltung/was-hilft-gegen-erkaeltung/<br>https://medizin-transparent.at/kochsalzloesung-nase/<br>https://www.meinmed.at/gesundheit/erkaeltung-hausmittel/1963<br>https://www.derstandard.at/story/1254311589371/troepfcheninfektion-grippeausbreitung-durch-kochsalzinhalation-verringern<br>https://www.sozialversicherung.at/cdscontent/?contentid=10007.862784 | SNI, drops or "inhalieren" is also supported by doctor's associations, such as Netdokter, Medicin transparant, MeinMed, and pharmacies , as well as older press articles |
| | ***Qualitative Research Summary - Austria*:**<br>see Germany, reviewed together with Germany | |
| **Belgium**<br>(Dutch)<br>CBIP (Fr)<br>= BCFI (Nl)<br><br>AB message: yes<br><br><br>Self medication: no<br>↓<br>↓<br>↓<br>Discussed: Decongestants | No official eHealth site targeting consumers<br>CBIP (Fr)= BCFI (Nl) = a repertory of medicines -> focus goes on<br>**11.5.2.11. Verkoudheid (Common cold)**<br><br>**Antibiotics are not indicated for colds (GRADE 1A).**<br>Nor is there currently an antiviral drug available that has been shown to prevent colds and/or complications or to lead to a faster return to daily activities. Consequently, there is no indication to use antiviral agents (GRADE 1C*).<br><br>https://www.cbip.be/fr/articles/query?number=F40F10B<br>Decongestants for nasal congestion (article dating from 2013)<br>• For the symptomatic treatment of **nasal congestion, vasoconstrictors** are sometimes administered orally or nasally. Their use should be done with caution, especially with regard to oral vasoconstrictors, due to potentially serious cardiac and/or neurological adverse effects. When administered nasally, problems such as drug-induced rhinitis or overdose may occur, especially with chronic use. In addition, caution is generally required in children, the elderly, people suffering from cardiovascular problems and during pregnancy. Drug combinations have an unfavourable benefit/risk ratio, except in certain patients with allergic rhinitis.<br>The site only starts from the different classes of medications.<br>For the site of ANSM: see France. | CIBP: Focus on decongestants. At the end of the page 'after lots of information on vasoconstrictors', the CIBP refers to old sources (links not valid anymore), such as :<br>• A link to ANSM = Agence Nationale de Sécurité du Médicament et des Produits de Santé, not of Belgium, but of France, yet this agency does not treat the subject of common cold, but many other diseases and 'dossiers thématiques" (see France), while upon searching on 'rhume' there appear only links to decongestant use.<br>• Vademecum Huisartsen 2000 – Today a book to be paid for and linking also to Domus Medica, yet where you have to have a log-in and have to be a member of the organization |

| **Country +** sequence elements from algorithm | **Relevant retrieved sections & Qualitative Research Summary**<br>- Structure of the e-site<br>- Section Treatment, self-care: content translated into English<br>Other links to supportive e-info, if applicable | **Comments on Internet Search**<br>- Sequence of sites popping up<br>- Major (official) eHealth site<br>- Links to other supportive sources |
|---|---|---|
| **Belgium**<br><br>Afmps= Fagg<br><br>↓<br>↓<br>↓<br>Only end of page: | Afmps. Conseils pour un bon usage des médicaments vasoconstricteurs par voie nasale utilisés en cas de rhume:<br>• **When and how to use vasoconstrictors (= decongestants) for nasal use**? .../...<br>• **Recommandations particulières** .../... lots of information on decongestants<br>• **Petit rappel : qu'est-ce que le rhume ?** = Information on common cold<br>• **[**Only at end of the long eHealth page**]: Que faire en cas de rhume ? (What to do when having a cold)**<br><br>[Content end of page]<br>First of all, it is recommended to:<br>• rinse your nose with saline solution;<br>• drink enough water;<br>• raise the head of the bed;<br>• avoid irritating fumes;<br>• renew and sufficiently humidify the air in the house.<br>Yet, the above points are subsequently followed by a big frame, which (confusingly) lists decongestant medicines 'médicaments concernés'<br><br>Only when you introduce the word "sérum physiologique" you find the instructions for children:<br>https://www.afmps.be/sites/default/files/downloads/afmps_MedicamentsEtEnfants_TouxRhume_Brochure.pdf | • Federal Agency for Medicines and health pops up (=Afmps in French, Fagg in Dutch).<br>• First focus on decongestants. Only at end of page information with the option for saline<br><br>• References go to other articles such as for children, yet is communication-wise, also ineffective, the saline option stated between the lines as obligatory before decongestant AFMPS, *Médicaments utilisés chez les enfants contre la toux et le rhume : recommandations de l'afmps*, https://www.afmps.be/fr/news/news_antitussifs_2013_04.<br><br>• To note: the articles also exist in Dutch, yet the Fagg (Dutch counterpart of Afmps) and none of the articles popped up on the screen upon searching. |
| | ***Qualitative Research Summary - Belgium:***<br>Belgium has no persuasive e-site targeting the patients seeking information.<br>• Of the e-sites visited, including French sites popping up, the focus is largely on (discouraging) decongestants.<br>• Only one site, originating from the Federal Agency for Medicines (AFMPS, Fagg) recommended SNI among the remedies to use for a common cold, located at the end of an e-page focussing on correct use of decongestants.<br>A recent check on eHealth messages about ABs, automatically proposed a flyer (on screen referred to by Gemini AI)[ 3]:<br>• this flyer used the same approach and slogan as in France, yet was coordinated by another commission (BAPCOC) [4].<br>• Both the flyer and organisation had not popped up during prior internet searches.<br>• The flyer lists many different indications for ABs, and recommends painkillers and antipyretics, yet no saline, for treatment (utmost steaming for sinusitis). | • |
| **Denmark**<br><br><br><br><br><br><br><br>AB message: yes | Sundhed.dk<br>Forkølelse<br>https://www.sundhed.dk/borger/patienthaandbogen/infektioner/sygdomme/virusinfektioner/forkoelelse/<br>• Hvad er forkølelse?<br>• Hvad er symptomerne på forkølelse?<br>• Hvordan stilles diagnosen?<br>• Hvorfor bliver man forkølet?<br>Hvordan behandler man forkølelse? [How to treat a cold?]<br>• Colds go away on their own. There is no medicine that can prevent, cure or shorten colds.<br>• **Antibiotics do not work against colds and are therefore not used.** Antibiotics can cause unnecessary side effects such as diarrhoea, fungal attacks on the skin and mucous membranes, development of resistance in bacteria in the environment, and are also a waste of money**.** | Sundhedsstyrelsen<br>= Danish Health Authority is the supreme authority of healthcare in Denmark.<br>The site just tells the disease cannot be treated, but the symptoms can be alleviated. The e-site does however not say how and continues with "complications", proposing antibiotics for bacterial infections. There is a lot of disease mongering.<br><br>Sundhed.dk = official eHealth site of Denmark<br><br>No other sites recommending saline for common cold were found, such as Netdokter.dk |

| **Country +**<br>sequence elements from algorithm | **Relevant retrieved sections & Qualitative Research Summary**<br>- Structure of the e-site<br>- Section Treatment, self-care: content translated into English<br>Other links to supportive e-info, if applicable | **Comments on Internet Search**<br>- Sequence of sites popping up<br>- Major (official) eHealth site<br>- Links to other supportive sources |
|---|---|---|
| Treatment:<br>↓<br>**Decongestant first-line**<br>↓<br>↓<br>↓<br>↓<br>↓<br>↓<br>Remedies<br>↓<br>↓<br>↓<br>↓<br>↓<br>↓<br>↓<br>↓<br>↓<br>↓<br>↓<br>↓<br>↓<br>↓<br>No vaccine | • However, medication can be used to relieve the symptoms. The most used preparations are nasal drops, cough and expectorants.<br>Medicin [Medicine]<br>If necessary, you can take an over-the-counter pain reliever, e.g. paracetamol, to reduce headache and fever<br>**Nasal spray causes the blood vessels in the mucous membrane to constrict** and relieves breathing through the nose. Nasal drops can therefore improve sleep<br>Cough syrup and expectorants, on the other hand, do not have any definite beneficial effect, neither in children nor in adults<br>Prolonged coughing in children raises the suspicion of asthma. For that reason, many have recommended using asthma medications to treat colds. However, treatment of long-term cough, which was not due to asthma, asthma medication was without effect compared to so-called placebo ("cheat medication").<br>Hvad kan jeg selv gøre? [What can I do?]<br>• Drink plenty of fluids - e.g. water or juice. This prevents you from becoming dehydrated<br>• Rest as much as you feel the need for it, even if rest is not a condition for recovery<br>• When you lie down, it can be a good idea to have a large pillow under your head; this usually reduces the tightness in the nose<br>• Avoid physical and mental stress as much as possible<br>• If you exercise regularly, you can try to exercise even if you have a cold. If it gets worse, of course you have to stop.<br>Alternativ medicin<br>• S Echinacea. Echinacea products are not effective - either for treating or preventing colds<br>• Taking vitamin C is not recommended<br>• Zinc - not recommended. Studies have not shown a clear effect<br>• Ask your doctor about coughs<br>Kindergarten, nursery and day care<br>Children in kindergarten, nursery and day care should stay at home if they have a fever above 38 degrees, or if they are involved to a degree that they cannot participate in the regular activities in the institution.<br>How do I avoid getting or worsening a cold?<br>• Hand hygiene. Infection mainly takes place indirectly through contact with viruses on surfaces. Therefore, hand hygiene is very important. Both hand sanitizer, water and soap neutralize the cold virus.<br>• Of course, you should also avoid close physical contact with people with a cold as far as possible to prevent direct infection.<br>• **Vaccination**<br>The large number of viruses means that it is practically impossible to develop a vaccine.<br>How does the disease develop?<br>The vast majority recover completely on their own without any other complications. In some cases, however, a cold can lead to strep throat, sinusitis, otitis media or pneumonia.<br>How widespread/frequent are colds?<br>• Almost the entire population is affected by a cold one or more times a year<br>• Children can have several colds a year. Adults typically have 2-4 colds a year<br>• Children aged ½-3 years are particularly often affected<br>• Colds are one of the most common reasons for visiting the doctor<br>Do you want to know more?<br>○ Sinusitis<br>○ Otitis media<br>○ Sore throat<br>○ Pneumonia | |

| **Country +** sequence elements from algorithm | **Relevant retrieved sections & Qualitative Research Summary**<br>- Structure of the e-site<br>- Section Treatment, self-care: content translated into English<br>Other links to supportive e-info, if applicable | **Comments on Internet Search**<br>- Sequence of sites popping up<br>- Major (official) eHealth site<br>- Links to other supportive sources |
|---|---|---|
| Physicians:<br>↓<br>↓<br>**Decongestants 1st-line** | ○ Cold - for healthcare professionals<br>○ **Antibiotics for respiratory infection** \| Choose Wisely (vaelgklogt.dk)<br><br>**Netdokter.dk**<br>**How can I relieve a cold?**<br>• If you don't have anything other than a cold, and the cold goes away within one to two weeks, there is no need to see a doctor.<br>• **Nasal agents to reduce swelling of the nasal mucosa can make breathing easier.**<br>• Sleep with your head up. With small children, you can lift the headboard of their bed, for example with a few phone books under the legs at the head of the bed.<br>• Avoid smoking, it disturbs the nasal mucosa.<br>• .../... at the end:<br>**How is a cold treated?**<br>• Colds are caused by viruses and are therefore not treated with antibiotics. It is only if complications arise, such as eye inflammation, sinusitis, otitis media and pneumonia, that medication may become necessary.<br>• If you are very bothered by a stuffy nose, you can use a nasal spray for a short period, which has an astringent effect on the nasal mucosa. | |
| | ***Qualitative Research Summary - Denmark***<br>The official eHealth sites rather proposed medicines to relieve common colds first-line, with a clearcut first-line recommendation for decongestants | |
| **Estonia**<br><br><br><br><br><br><br><br><br>AB message:<br><br><br><br><br><br><br><br><br><br>Treatment:<br>↓<br>**Decongestant first-line** | Meditsiiniportaal arst.ee<br>(Medical portal)<br>https://arst.ee/haigused-ja-seisundid/allergiline-nohu/<br>***Non-allergic rhinitis***<br><br>Rhinitis (runny nose) is an inflammation of the nasal mucosa, resulting in nasal congestion, nasal congestion and, depending on the etiology, accompanying symptoms (e.g. itching, sneezing, watery or purulent nasal congestion, anosmia (inability to smell)). Rhinitis is classified as allergic or non-allergic. Non-allergic rhinitis is usually caused by a viral infection, although irritants can also cause it. Diagnosis is usually clinical. Treatment includes humidification of room air, use of sympathomimetic amines and antihistamines. **Bacterial superinfection requires appropriate antibiotic therapy.**<br><br>There are several forms of non-allergic rhinitis.<br>(See also allergic rhinitis.)<br>• A bad cold: an acute runny nose, manifested by swelling and vasodilatation of the nasal mucosa, rhinorrhoea (runny nose) and obstruction (closure, obstruction, narrowing), is usually the result of a runny nose; **other causes include streptococcal, pneumococcal, and staphylococcal infections.**<br>• Chronic cold ..../....<br><br>***Treatment of non-allergic rhinitis***<br>For viral colds, decongestants, antihistamines, or both are used.<br>In case of atrophic rhinitis, topically with antibiotics, estrogens and vitamins A and D.<br>For vasomotor rhinitis, irrigation and sometimes topical corticosteroids and oral pseudoephedrine. | Health Authority: KRIIS.EE (zero hits)<br>https://www.terviseamet.ee/en<br><br>Not first-line supported by physicians' association Arst.ee<br>= A medical website that connects patients and doctors and introduces health products.<br>• Yet, saline is mentioned as the last option at a long information page, being called "often overlooked"<br><br>Also highly supported by brochure coordinated by:<br>Estonian Society of Infectious Diseases<br>Estonian Ambulance Association<br>Estonian Association of Pediatricians<br>Association of Family Nurses of the Estonian Nurses' Union<br>https://rakveretervisekeskus.ee/storage/app/media/hingamisteedea53trykk-1.pdf |

| **Country +** sequence elements from algorithm | **Relevant retrieved sections & Qualitative Research Summary**<br>- Structure of the e-site<br>- Section Treatment, self-care: content translated into English<br>Other links to supportive e-info, if applicable | **Comments on Internet Search**<br>- Sequence of sites popping up<br>- Major (official) eHealth site<br>- Links to other supportive sources |
|---|---|---|
| ↓<br>↓<br>↓<br>↓<br>AB message:<br>↓<br>↓<br>↓<br>↓<br>↓<br>↓<br>↓<br>↓<br>↓<br>↓<br>↓<br>↓<br>↓<br>↓<br>↓<br>↓<br><br>Saline last-line at the end of a long page | Viral colds can be treated symptomatically with decongestants (either topical vasoconstriction with a sympathomimetic amine such as oxymetazoline every 8-12 hours or phenylephrine 0.25% every 3-4 hours for no more than 7 days or systemic sympathomimetic amines such as pseudoephedrine 30 mg orally every 4 - after 6 hours). Antihistamines can be helpful, but drugs with anticholinergic properties dry out the mucosa and can therefore increase irritation. Decongestants can also relieve symptoms of acute bacterial common cold and chronic common cold, while the underlying bacterial infection requires culture (microbiological culture), identification of the pathogen, antibiotic susceptibility, and appropriate antimicrobial therapy. If symptoms persist, a biopsy may be necessary to rule out cancer.<br><br>Treatment of atrophic rhinitis …<br><br>Treatment of vasomotor rhinitis …<br>…/…<br><br>The most effective first-line drugs are:<br>nasal corticosteroids with or without oral or nasal antihistamines oral antihistamines plus oral decongestants (eg, a sympathomimetic such as pseudoephedrine).<br>Less effective alternatives are nasal mastoid cell stabilizers (eg, cromolyn) given 3 or 4 times daily, the nasal H1 blocker azelastine 1 to 2 times daily, and nasal ipratropium 0.03% 2 times every 4 to 6 hours, which relieves nasal congestion.<br>Nasal medications are often preferred over oral medications because less of the medication is absorbed systemically.<br><br>*"Intranasal saline, often overlooked, helps mobilize thick nasal secretions and hydrate the nasal mucosa; various saline kits and flushing devices (eg, squeeze bottles, pump syringes) are commercially available, or patients can prepare their own solutions*."<br><br>Yet, saline is highly supported in a brochure online, supported by a large number of Medical Associations: https://rakveretervisekeskus.ee/storage/app/media/hingamisteedea53trykk-1.pdf | |
| | ***Qualitative Research Summary - Estonia***<br>SNI is also traditional to some extent, the official eHealth sites rather proposed medicines to relieve common colds first-line, with a clearcut first-line recommendation for decongestants | |
| **Finland**<br>Self-treatment?<br><br><br><br><br><br><br><br><br><br>Self-treatment<br>↓<br>↓<br>↓ | Flunssa, nuhakuume: https://www.terveyskirjasto.fi/dlk00590 (translates to flu; yet influenza = Influenssa<br>See Box: ..//… "Symptoms can be alleviated with self-care" …//…<br>• **General: …//…**<br>• **Catching the flu: b…//…**<br>• **Diagnosing the flu (=cold):** …//…<br>• **When to contact a doctor?…//…**<br>Voit tehdä Omaolossa oirearvion …… You can do a symptom assessment in Omaolo if you have symptoms of inflammation in the nose, pharynx, larynx, sinuses, trachea or lungs. It is intended for assessing the need for treatment of all respiratory infection symptoms, including coronavirus disease. The survey helps you assess when and what kind of treatment you should seek, and how you can treat your symptoms yourself.<br><br>**Flunssan itsehoito = Self-treatment of the flu (=cold)**<br>Cold are caused by viruses and get better without treatment. Self-care focuses on alleviating symptoms.<br>• The primary treatment for flu symptoms is rest. Physical exertion should be avoided in order to avoid the development of possible complications. Light movement and tinkering are not a disadvantage. It is safe to | The site is based on Duodecim (Evidence-Based Medicine Guidelines): the e-site treats diseases such as flu (equivalent used for the common cold), influenza, evolution to pneumonia all under same denominator, then goes to 'when to see a doctor', before the treatment is addressed, where saline is mentioned between several medications listed, as is done with steaming. The strong point is that a link is given to how to irrigate.<br><br>The site THL (=THL is a national expert institute that offers reliable information to support decision-making and operations in the health and well-being sector) does not have the target word flunssa as a title, and 40 hits pop up that one has to search through. One gives instructions via flunssa over the flue, thereby proposing steam to alleviate cough, yet not saline, as Netherlands |

| **Country +** sequence elements from algorithm | **Relevant retrieved sections & Qualitative Research Summary**<br>- Structure of the e-site<br>- Section Treatment, self-care: content translated into English<br>Other links to supportive e-info, if applicable | **Comments on Internet Search**<br>- Sequence of sites popping up<br>- Major (official) eHealth site<br>- Links to other supportive sources |
|---|---|---|
| ↓<br>↓<br>↓<br>↓<br>↓<br>AB message<br>↓<br>Decongestant limited use<br>↓<br>**Use saline for relief + video**<br>↓<br>↓<br>Cough : medicines not to use<br>↓<br>↓<br>Other remedies, such as honey & Vaporub & steam<br>↓<br>↓<br>↓<br>Other remedies<br>↓<br>↓<br>↓<br>↓<br>↓<br>↓<br>↓<br>↓<br>↓<br>↓<br>↓<br>Hygiene measures<br>↓<br>↓<br>↓<br>↓<br>↓<br>↓<br>↓ | start exercising as much as you can when the general symptoms (fever and general aches) are over. This usually means a 3-5 day break from exercise.<br>• The fever and aches associated with the flu can be alleviated with anti-inflammatory drugs or paracetamol, depending on the judgement. Acetylsalicylic acid (ASA, aspirin) is not recommended as an antipyretic for children and adolescents; see Painkillers - safe use.<br>• **Antibiotics are not useful in the common cold. There is no cure for rhinitis, but its symptoms can be alleviated with medication.**<br>• Nasal congestion and discharge can be alleviated with nasal vasoconstrictor drugs (xylometazoline, oxymetazoline). The preparations should only be used for a short time (depending on the preparation and the age of the patient, for a maximum of 5–7 days).<br>• **Rinsing the nose with a nasal spray often relieves the symptoms. Watch a video on using a nasal canister here**<br>• **For children under one year of age, nasal drops or sprays containing physiological common salt can be used.** Tablets or capsules used for allergic rhinitis do not help nasal congestion caused by the flu.<br>• Cough is a protective reaction of the respiratory tract in connection with inflammation and irritation. Its function is to remove mucus. The routine use of cough suppressants to treat cough associated with the flu is not recommended. Based on research, cough medicines do not have a significant effect on acute cough symptoms and can have adverse effects. Children's coughs are not recommended to be treated with over-the-counter cough medicines, as their possible harms are greater than the possible benefits.<br>• Honey may be helpful for children's coughs. It can be used in children over 1 year old.<br>• Menthol ointment (Vicks VapoRub®) may help with a cough. It can be used in children over 2 years old.<br>• The best treatment for hoarseness (inflammation of the larynx) is to avoid talking. Whispering, however, strains the vocal cords more than speaking in a low voice.<br>• Steam inhalation can be helpful in the treatment of cough and congestion. Steaming the shower with lukewarm water may ease congestion and cough. Breathing in boiling water from the boiler poses a risk of burns and should definitely be avoided. Safe vaporizers are also available.<br>• The use of zinc does not reduce the number of infections, and the evidence for the effectiveness of the lozenge in shortening the symptoms is uncertain. Abdominal symptoms and a bad taste in the mouth may be associated with the use. Nasal sprays containing zinc should not be used. Zinc preparations should not be used in children.<br>• Continuous use of large doses of vitamin C does not prevent getting the flu, but it may slightly shorten the duration of the symptoms. Vitamin C does not seem to be useful for flu symptoms if its regular use is started only after getting the flu.<br>• It is necessary to take care of drinking enough, especially in the case of a small child and an old person with a fever.<br>• The need for sick leave is usually determined by general health and the nature of the work. Usually 1–3 days of sick leave is enough.<br><br>Flu prevention<br>• Flu viruses are effectively transmitted by hand contact. The most important way to prevent the flu is careful hand hygiene. It means washing your hands with soap and water or disinfecting your hands with a product that kills viruses. Hands should be washed at least when entering from outside, after using the toilet and before eating, and after coughing, sneezing or coughing.<br>• Coughing into a handkerchief or your own sleeve is recommended when moving around people. The used handkerchief is immediately put in the trash.<br>• It is good to avoid touching the area of the mouth, nose, eyes and face, unless you have just washed your hands. | does for cough upon a common cold. Another link for rhinovirus does not specify the 'symptomatic' treatment<br><br>Current site however states under nasal irrigation:<br>https://www.duodecimlehti.fi/duo16327<br>"Although the evidence for the effectiveness of rinsing in connection with sudden infections is incomplete, rinsing could also be used during a common cold or a COVID-19 infection, as long as care is taken to clean the rinsing device and the environment."<br>…/…<br>Saline nasal irrigation refers to rinsing the nasal cavity with a physiological saline solution. Its mechanisms of action are not yet certain. It is assumed that at least the thinning and mechanical removal of the mucus layer, the reduction of swelling and antigen load, and the improvement of cilia clearance and cilia function of the nasal and side cavities are important (3,4). Rinsing may also remove stimuli that come with the "reat'Ing air, such as allergens, irritants, particles and inflammatory mediators (3,4). In addition, the effect may be partly based on the moistening of the nasal mucosa and better absorption of the local medicine (such as glucocorticoid) (3,4).<br>…/…<br>Implementation of saline nasal irrigation:<br>There are several rinsing devices on the market that work in different ways, which can be divided based on their mechanism of action into small volume and low pressure (salt sprays, nebulizers), large volume and low pressure (nasal can, pump bottle) and large volume and high pressure (trade names Neilmed, Sinugator and Navage), followed by lots of explanation!<br><br>Tapiala 2020 describes how popular nasal irrigation is in Finland across doctors and pharmacy practices :<br>the majority of the respondents in their study recommended nasal saline irrigation for their patients either as a symptomatic treatment (98.0%) or to treat a specific condition (97.5%), 68% recommending it for common cold, such as acute rhinosinusitis, but also for chronic rhinosinusitis and allergic rhinitis. Yet they tried to make doctors aware of potential 'adverse events' (epistaxis, pain, and dryness of the nose). |

| Country + sequence elements from algorithm | Relevant retrieved sections & Qualitative Research Summary<br>- Structure of the e-site<br>- Section Treatment, self-care: content translated into English<br>Other links to supportive e-info, if applicable | Comments on Internet Search<br>- Sequence of sites popping up<br>- Major (official) eHealth site<br>- Links to other supportive sources |
|---|---|---|
| ↓<br>↓<br>↓<br>↓<br>↓<br><br>No vaccine | • Continuous intake of large doses of vitamin C does not reduce the number of infections but may slightly shorten the duration of symptoms.<br>• Regular exercise does not reduce the number of infections but may alleviate their symptoms and shorten their duration.<br>• Vitamin D and probiotics, when used regularly, may slightly prevent respiratory infections.<br>• There is no vaccine for the flu (=the common cod), but there is for the flu (= influenza). | |
| | ***Qualitative analysis - Finland***<br>For Finland, the broad traditional use of SNI has been well described [2]; 69% of physicians and pharmacists were found to recommend SNI in this country. Finland was given the score 2, as the DUODECIM-based information of the Finnish Medical Society (our first hit) places SNI after proposing the decongestants for alleviation of the symptoms, yet limits their use by a short message; SNI is further proposed as often relieving symptoms yet with a link with instructions on how to prepare and how to do it, so overall communicating positively and persuasively on SNI. | |
| **France**<br>**ANSM**<br><br><br>No algorithm elements:<br>↓<br>↓<br>only info on supply chain, antibiotics inclusive<br><br><br><br><br><br><br>↓<br>↓<br>↓<br>↓<br>↓<br>Antivirals<br>↓<br>Vaccination<br><br>↓<br>↓<br>↓<br>↓<br>↓<br>↓<br>↓ | For the first French e-Health hits: see column on the right-hand side: all sites with lots of medicinal alternatives being proposed and disease mongering<br><br>= Agence nationale de sécurité du médicament et des produits de santé<br>https://ansm.sante.fr/actualites/a-la-une<br><br>[Content excerpts]:<br>**[First topic] "Plan hivernal" (winter plan) – translation of communication:**<br>https://ansm.sante.fr/dossiers-thematiques/plan-hivernal<br>We are deploying a winter plan, built with patient associations, representatives of healthcare professionals and all players in the supply chain. This plan aims to anticipate and limit tensions over certain major winter medications and thus secure their availability in order to meet patient needs.<br>We are stopping the weekly publication of winter plan indicators but we are maintaining close monitoring of supplies for these drugs, as we do for other treatments.<br>Fight against drug shortages: the ANSM activates its 2023-2024 winter plan<br>In this context, the ANSM carries out reinforced monitoring of data on the supplies of certain medicines (stocks and supplies of laboratories, wholesale distributors and pharmacies, monitoring of sales in pharmacies):<br>• Antibiotics<br>• Fever medications<br>• Corticosteroids administered orally<br>• Asthma Medications<br><br>**[2nd topic]. Le virus respiratoire syncytial (VRS) (Respiratory Syncytial Virus**<br>**…/…**<br><u>Immunization of infants against RSV</u><br>To protect infants from severe forms of bronchiolitis, two medications are currently available:<br>• Beyfortus (nirsevimab)<br>• Synagis (palivizumab).<br>These two drugs are monoclonal antibodies which help strengthen the infant's immune system: the antibody is supplied directly to the body and helps it defend itself against RSV.<br><br>The indications of Beyfortus and Synagis are, however, different. | First target word leads to:<br>- **<u>VIDAL (ABC of medicines)</u>**<br>https://www.vidal.fr/maladies/nez-gorge-oreilles/rhume-rhinite.html -> select 'Traitements':<br>• Les solutions de lavage nasal<br>• Les médicaments contre le rhume par voie nasale<br>○ Les solutions nasales antibactériennes et les inhalations<br>○ Des solutions nasales antibactériennes permettent de désinfecter le nez.<br>• Les solutions nasales décongestionnantes<br>• Les solutions nasales anti-inflammatoires<br>• Les médicaments contre le rhume par voie orale<br>• Les autres médicaments<br><br>- **<u>Passeport Santé</u>** :<br>400 hits for 'rhume' -> first hit<br>https://www.passeportsante.net/https://www.passeportsante.net/fr/Maux/Problemes/Fiche.aspx?doc=rhume_pm<br>• Rhume<br>• Symptômes et facteurs de risque<br>• Prévention<br>• Traitements médicaux: broad list of medications, including decongestants, anti-inflammatory, Zinc….<br>• L'opinion de notre médecin<br>• Approches complémentaires<br>[No saline]<br>- **MSD Manuels** : a pharma-site of MSD proposing medicines |

| **Country +** sequence elements from algorithm | **Relevant retrieved sections & Qualitative Research Summary**<br>- Structure of the e-site<br>- Section Treatment, self-care: content translated into English<br>Other links to supportive e-info, if applicable | **Comments on Internet Search**<br>- Sequence of sites popping up<br>- Major (official) eHealth site<br>- Links to other supportive sources |
|---|---|---|
| ↓<br>↓<br><br><br><br>Saline for children in ‘bronchilolitis’<br><br><br><br><br><br><br><br><br><br>Warnings against decongestant use:<br><br><br><br><br>**Decongestants**<br>↓<br>↓<br>↓<br>↓<br>↓<br>↓<br>↓<br>↓<br>↓<br>↓<br>↓<br>↓<br>↓<br>↓<br>↓<br>↓<br>↓<br>↓<br>↓<br>↓<br>Saline | • Beyfortus is indicated for the prevention of lower respiratory tract infections due to RSV in neonates and infants during their first season of RSV circulation.<br>• Synagis is indicated for the prevention of serious lower respiratory tract infections due to RSV, requiring hospitalization in certain children at high risk of RSV infection.<br><br>Management of bronchiolitis<br>The treatment is essentially based on ***“lavage” of the child's nose*** and monitoring the evolution of their state of health:<br>Certain signs require an appointment with the child's doctor: change in behavior, faster breathing, hollowing of the chest, infant drinking less for several meals<br>Other signs require a call to 15: cyanosis (bluish discoloration) around the mouth, discomfort, breathing pauses, infant sleeping all the time.<br><br>**[Indirect link]“En cas de rhume, évitez les médicaments vasoconstricteurs par voie orale !”**<br>https://ansm.sante.fr/actualites/en-cas-de-rhume-evitez-les-medicaments-vasoconstricteurs-par-voie-orale<br><br>[Repeated internet access: 24.06.2024]<br>Only when combining with the word “sérum physiologique”, you find the page on:<br>https://ansm.sante.fr/actualites/rhume-nez-qui-coule-nez-bouche-attention-lutilisation-des-vasoconstricteurs-expose-a-des-risques-soyez-vigilants<br>“**Rhume, nez qui coule, nez bouché** ? Attention : l’utilisation des vasoconstricteurs expose à des risques, soyez vigilants !”<br>Lengthy page on decongestant use ->The focus first goes to decongestants, you shouldn’t use.<br>• If you want to take a vasoconstrictor<br>- Inform your pharmacist of your medical history: he will be able to tell you if you can take this treatment.<br>- Always follow the dosage (dose and frequency of intake) recommended by your pharmacist.<br>- Do not exceed 5 days of treatment: consult your doctor if your symptoms persist or worsen.<br>- Do not combine these medicines with another vasoconstrictor taken orally or nasally or another medicine containing paracetamol, ibuprofen or an antihistamine. The composition of the medicine is indicated on the main side of the box.<br>- Do not use any of these medicines in children under 15 years of age.<br>- Vasoconstrictors are strongly discouraged throughout pregnancy and should never be used from the end of the 5th month of pregnancy when they include ibuprofen. Always seek the advice of a doctor or pharmacist before taking these medications if you are pregnant.<br>- If breastfeeding, vasoconstrictors are strictly prohibited.<br>• If you have taken a vasoconstrictor<br>Pay attention to symptoms that may indicate the occurrence of a stroke or myocardial infarction (for information, see the patient information document). If any of these symptoms appear, stop taking this medication immediately and consult your doctor immediately. Be careful, these adverse effects can occur regardless of the dose and duration of exposure to these medications.<br>• Followed by (a link to) : “Helping (instruction) sheet for dispensing oral vasoconstrictors” (updated version of November 2022)<br>The recommendations on what to do (saline first-line) only appears after scrolling down under a heading<br>• **As a reminder, colds disappear spontaneously after 7 to 10 days without treatment.**<br>While waiting for spontaneous healing, several comfort solutions can be adopted: | https://www.msdmanuals.com/<br>[Recommendation of saline, if any, is lost among other medicines actively offered and complications/ disease mongering ]<br><br>**AMS:**<br>The official French agency does not treat the subject of common cold, but many other diseases under the heading ‘Domaine Médical’ and they do not treat non-pharmacological approach under ‘Produit de Santé’, listing “Médicaments”, “Dispositifs médicaux”, “Dispositifs médicaux de diagnostic in vitro”, “Produits biologiques”, “Vaccins”, “Stupéfiants & psychotropes”, “Cosmétiques”<br><br>• They also shows ‘Dossiers Thématiques” , one corresponding to a sneezing woman with common cold. Opening the site, there are 2 topics:<br>• Winter plan: contains only information on drugs, antibiotics in particular<br><br>• 2nd topic: RSV<br>After information on the disease, follows first immunisation – with links to the different vaccines and vaccines to come – this before the section on how to treat (=end of the communication) – No explanation on how to perform nasal lavage<br><br>• [Indirect links identified by searching on ‘lavage’ and ‘saline’ ] Only when searching on “lavage” a link arrives to the following topic, where lavage nasale are recommended first line<br><br>Only after seeking for “Santé information France “ and adding the word “officiel” (= official) [accessed 10.08.2024], we find articles under an official site of the government: Santé.fr : 2492 hits!<br><br>The first link treats ‘Rhinopharyngite ou Rhume”<br>https://www.sante.fr/antibiomalin-pour-savoir-comment-bien-utiliser-les-antibiotiques/rhinopharyngite-ou-rhume<br><br>The option ‘serum physiologique ‘is hidden between other lines. No information on how to prepare. There is much more focus on the use of antibiotics and advise on non-pharmacological interventions such as regular handwashing, using alcohol-based antiseptics, coughing in the elbow, avoid contact and wearing masks. |

| **Country +** sequence elements from algorithm | **Relevant retrieved sections & Qualitative Research Summary**<br>- Structure of the e-site<br>- Section Treatment, self-care: content translated into English<br>Other links to supportive e-info, if applicable | **Comments on Internet Search**<br>- Sequence of sites popping up<br>- Major (official) eHealth site<br>- Links to other supportive sources |
| --- | --- | --- |
| ---------------------<br>Santé.fr<br><br>Page on: Rhinopharyngite ou rhume<br><br>**AB message + video's**<br><br>…//…<br><br>↓<br>↓ | - moisten the inside of your nose with suitable washing solutions: physiological serum, thermal water or sea water sprays, etc.<br>- drink enough<br>- sleep with your head elevated<br>- make sure to maintain a cool atmosphere (18-20°C maximum) and ventilate the rooms regularly<br>You find through links , also a fiche:<br>https://ansm.sante.fr/uploads/2022/12/14/20221214-livret-patient-vc-nov-2022.pdf<br>The "solutions de comfort " (saline to moisturize), is followed by ample information on decongestants, a.o. claiming "if you want to take a medicine against cold symptoms "->->Saline is proposed little convincing as a "comfort solution" to moisten yet not proposed to relieve the cold or its symptoms<br><br>Only after seeking for "Santé information France " and adding the word "officiel" (= official) [accessed 10.08.2024], we find articles under an official site of the government: Santé.fr : 2492 hits!<br><br>The first link treats**:**<br>**"Rhinopharyngite ou Rhume"**<br>https://www.sante.fr/antibiomalin-pour-savoir-comment-bien-utiliser-les-antibiotiques/rhinopharyngite-ou-rhume<br>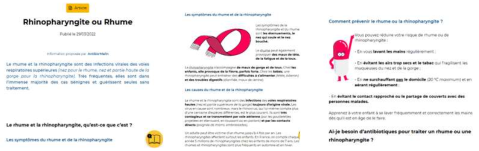<br>**Do I need antibiotics to treat a cold or nasopharyngitis?**<br>Colds and nasopharyngitis are always of viral origin: antibiotics are of no use. The use of antibiotics for viral infections exposes you to unnecessary risks of frequent side effects (digestive disorders, fungal infections, etc.) and potentially serious ones.<br><br>Videolinks on ABs by 'Antibiomalin' (pill-looking creature):<br>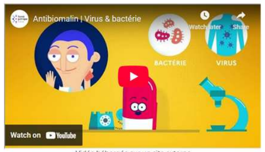<br>Vidéo hébergée sur un site externe<br><br>**Les antibiotiques sont-ils efficaces sur certaines maladies hivernales ?**<br>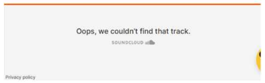 | While saline is a recommendation in the second hit (https://www.sante.fr/rhume-rhinite ) , it simply did never pop up during our initial searches of the Internet and was detected only at the end of editing our article. Moreover, no self-preparation is explained, but the use is guided to commercial formulations, even uni-doses (=single doses), available in the pharmacy. Immediately, other medicinal options (disinfectant, or medicines relieving symptoms) are proposed that therefore may be preferred by consumers.<br><br>The youtube videos concerning avoiding ABC ("Antibio" – "Antibiomalin") are comics and ends with the message that "les antibiotiques: bien se soigner, c"est d'abord bien les utiliser" (=Antibiotics: good self-care means using them properly first), rather than saying they are generally not needed to treat common colds. This message is also put up on the header of this site, and so – unwillingly – creates need for using antibiotics rather than immediately discouraging it. |

| **Country +** sequence elements from algorithm | **Relevant retrieved sections & Qualitative Research Summary**<br>- Structure of the e-site<br>- Section Treatment, self-care: content translated into English<br>Other links to supportive e-info, if applicable | **Comments on Internet Search**<br>- Sequence of sites popping up<br>- Major (official) eHealth site<br>- Links to other supportive sources |
|---|---|---|
| Self-treatment<br>↓<br>↓<br>↓<br><br>**Only for child:**<br><br>Saline ( for child)<br><br>-------------------<br>Santé.fr<br><br>eHealth page on: “cold rhinitis”<br>Saline single doses recommended<br>+ Local **antibacterial** treatments recommended<br>↓<br>**decongestant** | **Combien de temps vais-je souffrir d'un rhume ou d'une rhinopharyngite ? Quand aller chez le médecin ?**<br><br>**Les rhumes guérissent spontanément, sans traitement, en 7 à 10 jours.**<br><br>L**es rhinopharyngites guérissent également seules dans la quasi-totalit des cas.** L'écoulement nasal et la toux peuvent toutefois persister **10 à 15 jours.**<br><br>Les changements de couleur des sécrétions du nez (transparentes, jaunes, vertes...) font partie de l'évolution naturelle de la maladie. Ils ne sont pas du la présence d'une surinfection par une bactérie.<br><br>**En cas de fièvre persistant au-delà de 3 jours, une surinfection de type otite ou sinusite peut être en cause :** consultez votre médecin.<br><br>**Que puis-je faire pour soulager les symptômes du rhume ou de la rhinopharyngite ?**<br><br>**What can I do to relieve cold or nasopharyngitis symptoms?**<br>Rest if possible and drink enough, at least 1.5 liters of water per day. Make sure to maintain a cool atmosphere (between 18 and 20°C) in your home, particularly in the bedrooms. Sleep with your head elevated to reduce nasal congestion. Try to stop smoking and avoid smoky environments.<br><br>If your child has a fever, don't cover him too much and give him something to drink regularly, even if he doesn't cry out.<br>You can take paracetamol (or give it to your child in a dosage appropriate to their weight) to relieve fever, sore throat or headaches. Ibuprofen should be avoided as it can increase the risk of complications. Washing the nose with physiological serum or with a spray of sea or thermal water can help clear nasal obstruction. There are products suitable for children: ask your pharmacist for advice.<br><br>**How to protect my environment?**<br>Wash your hands regularly or use a hydroalcoholic solution. Use single-use tissues and cough into your elbow. As long as you are sick, avoid contact with vulnerable people (infants, elderly people, immunocompromised patients) and, if necessary, wear a mask.<br><br>“The information contained in this sheet is general and does not replace the advice of your healthcare professional.”<br><br>->-> The option ‘serum physiologique ‘ only pops up at the complete end of the e-Health article, after scrolling – we initially failed to see it : a consumer may not have scrolled up to, staying and leaving the page after having seen the youtube video on use of antibiotics. The recommendation is hidden between other lines. No information on how to prepare. There is much more focus on the non-pharmacological messages such as regular handwashing, using alcohol-based antiseptics, coughing in the elbow, avoid contact and wearing masks. Next (second) hit on the e-Site allows saline to pop up first line, yet immediately followed by the option to use nasal antibacterial treatments.<br><br>**The second hit on the same e-Health site: “rhume (rhinites)”**<br>This page recommends saline, yet immediately also offers other options that may look more attractive to consumers” (https://www.sante.fr/rhume-rhinite ):<br><br>**How to treat a cold?**<br>• Physiological serum or sea water solutions, sold in pharmacies and drugstores, allow you to wash and unclog the nose effectively; prefer products packaged in single doses.<br>• Local antibacterial treatments also make it possible to disinfect the nose and inhalations to clear the nose. |  |

| **Country +** sequence elements from algorithm | **Relevant retrieved sections & Qualitative Research Summary**<br>- Structure of the e-site<br>- Section Treatment, self-care: content translated into English<br>Other links to supportive e-info, if applicable | **Comments on Internet Search**<br>- Sequence of sites popping up<br>- Major (official) eHealth site<br>- Links to other supportive sources |
|---|---|---|
| | • Medicines in the form of tablets or capsules are offered to relieve cold symptoms if they are too bothersome. They may contain paracetamol or ibuprofen to combat fever and headaches, an antihistamine which has a drying effect on mucus secretion or a vasoconstrictor which has a decongestant effect giving the sensation of breathing more freely. Although these medications are sold without a prescription, their use requires the greatest caution:<br>• check contraindications (children under 15 years old, pregnancy, heart problems, combination with other medications, etc.) with your pharmacist;<br>• do not exceed the daily dose recommended by your pharmacist;<br>• do not extend treatment beyond 5 days. | |
|  | ***Qualitative Research Summary - France***<br>For France, there was no immediate access to a well-structured public health page on the Common Cold, searching for "le rhume", but rather a confusing labyrinth of fractionated messages over different e-sites.<br>• The first hit which first popped up was governmental site ANSM [5], revealing only information on missing medications due to a shortage of their delivery to pharmacies, and on the prevention of the flue (influenza) by vaccination.<br>• The first following relevant hit was VIDAL [6], the major French library for medications, both for selfcare and professionals.<br>• Yet, while proposing first-line "le sérum physiologique" (= isotonic saline) for common colds, this site overemphasizes the many medicinal options - from simple alternatives, such as nasal antiseptic or other medicinal sprays, to decongestants and antivirals - in such a way that the first-line recommendation of SNI is lost and that patients may prefer a medicine.<br>• This is also effectuated by the extensive documentation of complications, whereby ABs may be needed: hence, serious disease mongering is created, known in internet promotion to typically drive the readers to medicate.<br>The next e-pages popping up also originated from the ANSM, one yet focusing on the restriction of oral decongestants [7], another on RSV/bronchiolitis [8], where 'lavage' of saline is only proposed at the far end of the page 'how to protect' after information on vaccination in pregnant women and prevention with monoclonal antibodies in infants.<br>• Only when also searching for "sérum physiologique" combined with the topic for this site, information targeted to 'le rhume' appeared, yet again with first-line lengthy focus on restriction of decongestants, without antibiotic message, and saline only proposed at the very end of the page, just for moistening the nose [9].<br>Only during a check for the AB message, adding the target word "antibiotics", the site "Santé.fr" popped up, appearing to be the official French eHealth site, yet only publishing numerous articles, rather than being a well-structured instructive e-site for selfcare of 'le rhume' (the common language used by the public):<br>• the two first articles of 14 hits for the target word "rhume" were entitled "Rhinopharyngite ou Rhume" [10] and "Rhume/rhinite"[11]: they included recommendations of saline, yet these articles never popped up during our initial searches of the Internet, while the option to use saline was moreover first missed during the initial analysis: its recommendation remained unnoticed between other sentences, below a video, and/or were rather linked to child treatment or "moistening the nose", not proposed for relief. |  |

| **Country +** sequence elements from algorithm | **Relevant retrieved sections & Qualitative Research Summary**<br>- Structure of the e-site<br>- Section Treatment, self-care: content translated into English<br>Other links to supportive e-info, if applicable | **Comments on Internet Search**<br>- Sequence of sites popping up<br>- Major (official) eHealth site<br>- Links to other supportive sources |
|---|---|---|
| | • On the page 'rhinite' [11], the proposition of saline or seawater for washing and unclogging the nose - sold in pharmacies and drugstores and recommending 'unit' doses - was immediately followed by the proposition of "local antibacterial treatments making it possible to disinfect the nose", so promoting an antimicrobial concept, therefore possibly fuelling misbeliefs, and sounding more promising than nasal saline. So, overall SNI was lost between the many options and messages.<br>• The message on ABs was only found on the "Rhinopharyngite" page [10] linking the messages to YouTube videos, introducing "Antibio Malin" (a pill-like creature called "Smart AB"): in these video's, the speaking pill-creature proposes situations where (or where not) to use ABs. These videos deviate attention from the common cold page: they seem little viewed online, so questioning their impact on patients' attitudes.<br>• The message is: "*Les antibiotiques: bien se soigner, c"est d'abord bien les utiliser*" (=Antibiotics: good self-care means using them properly first) – a message seemingly confusing when reading further under treatment: "*Local antibacterial treatments also make it possible to disinfect the nose and inhalations to clear the nose".* This e-page appeared to be a subsection from the e-pages on antimicrobial resistance, also carrying this slogan. | |
| **Germany**<br><br><br><br><br><br><br><br><br><br><br>Prevention (Vitamins, Echinacea, probiotics)<br>↓<br>Treatment<br>↓<br>↓<br>↓<br>Decongestant<br>↓<br>AB message<br>↓<br>Other: zinc<br>Vit D<br>+ (herbal) remedies | https://www.gesundheitsinformation.de/erkaeltung.html<br>Erkältung = Common Cold:<br>• Einleitung<br>• Symptome<br>• Ursachen<br>• Häufigkeit<br>• Verlauf<br>• Folgen<br>• Diagnose<br>• Vorbeugung = Prevention<br>Sometimes vitamins or echinacea (preparations containing **echinacea extracts**) are advertised as preventing colds. Some people start taking these remedies a few weeks before cold season. However, their protective effect is mostly limited. However, studies provide evidence that **probiotics** can help reduce the risk of colds - especially upper respiratory tract infections. If you have a vitamin D deficiency, vitamin D supplements may also help prevent colds.<br>• Behandlung = Treatment<br>• To date, there is no medication that can specifically combat cold viruses and noticeably shorten the duration of the illness. This is because there are so many different cold viruses. However, a number of **remedies** can alleviate some of the symptoms somewhat. These include fever-reducing agents and painkillers such as ibuprofen, paracetamol or acetylsalicylic acid (ASS) as well as **decongestant nasal** sprays for short-term use.<br>• **Antibiotics** do not work against viruses and should only be used if bacteria are also involved and have led to complications. In addition, antibiotics often have side effects.<br>• Preparations with zinc, vitamin C or echinacea extracts are often recommended against colds. To date, the advantages and disadvantages of these agents cannot be reliably assessed because there is little study data or the studies show contradictory results. According to studies, vitamin D does not seem to be effective against colds. | Gesundheitsinformation.de has a similar layout as the one by Austria, yet differs considerably behind the link to more information, leading in this case to information on remedies such as vitamin C:<br>https://www.gesundheitsinformation.de/schuetzt-vitamin-c-vor-erkaeltungen.html<br><br>[` see also Austria] |

| **Country +** sequence elements from algorithm | **Relevant retrieved sections & Qualitative Research Summary**<br>- Structure of the e-site<br>- Section Treatment, self-care: content translated into English<br>Other links to supportive e-info, if applicable | **Comments on Internet Search**<br>- Sequence of sites popping up<br>- Major (official) eHealth site<br>- Links to other supportive sources |
|---|---|---|
| + Honey<br><br><br><br><br><br><br><br><br>Saline 1srt line Nasenpflege / inhalieren mit Kochsalz<br>↓<br>↓<br>↓<br>↓<br>↓<br>↓<br>↓<br>↓<br>↓<br>↓<br>…//… | • Honey or herbal medicines such as certain extracts of geranium root, primrose root, thyme, eucalyptus or ivy leaves may provide some cough relief<br><br>See also : https://www.gesundheitsinformation.de/schnupfen-husten-und-halsschmerzen-lindern.html<br><br>Yet, abundant e-health sites by doctors, hospitals, pharmacists, natural medicine, insurance companies and other associations, as well as Public Media, recommend Kochsalz Losung for a cold (Schupfen) (NaselPflege)<br><br>https://www.stiftung-gesundheitswissen.de/sites/default/files/pdf/2018_10_30_Infozepte_Inhalation_OnePage_v1_ts.pdf<br>https://www.hno-aerzte-im-netz.de/therapien/inhalieren.html<br>https://hno-johannssen.de/?page_id=74<br>https://www.netdoktor.de/therapien/inhalieren/mit-salz/<br>https://www.aok.de/pk/magazin/koerper-psyche/organe/richtig-inhalieren-mit-hausmitteln-und-medikamenten/<br>https://www.cegla.de/wissen/inhalation-mit-kochsalz/<br>https://www.zdf.de/nachrichten/ratgeber/gesundheit/inhalieren-mit-salz-bei-husten-100.html<br>https://www.ndr.de/ratgeber/gesundheit/Erkaeltung-und-Schnupfen-Diese-Hausmittel-helfen,erkaeltung196.html<br>https://www.apotheken-umschau.de/familie/kindergesundheit/kochsalz-nasentropfen-kleinkinder-schneller-durch-mit-der-erkaeltung-1165117.html<br>https://www.muenchener-verein.de/ratgeber/gesundheit/hausmittel-bei-erkaeltung/<br>…//….//….//…. | |
| | ***Qualitative analysis - Germany***<br>Germany and Austria were reviewed together: their e-sites share the same language, and both internet sites always popped up simultaneously on the computer screen during the internet searches per country domain.<br>• In Austria and Germany, "Nasenpflege" (care of the nose) with saline, or inhalation of saline, is a well-known popular, familiar tradition. This traditional use was confirmed by ample communications by German doctors (individual or associations), as well as other health care providers (see inventory in **Table S2**, e.g., HNO-Ärzte), and by a publication on selfcare in Austria [1].<br>• Yet, on the main page of their well-instructive eHealth sites (both similarly built up for access by the consumer), no references to SNI were found as a choice for the common cold (even not for children).<br>• On the German site, a link to a separate webpage with treatment options proposed decongestants, vitamin C and D, zinc, honey, steaming with or without substances such as Kamille or peppermint oil.<br>• The Austrian governmental eHealth site, in contrast, did not discuss, nor offer a link to a webpage with these medicated options: the concise eHealth page for the common cold deviated his readers for more information on treatment to the e-portal of the official Austrian Health Insurance, which communicates that there are no medicines that work for the common cold and proposes selfcare first-line, including SNI, before discussing medication/medicines [12]. To note, Austria has the lowest ABC. | |

| Country + sequence elements from algorithm | Relevant retrieved sections & Qualitative Research Summary<br>- Structure of the e-site<br>- Section Treatment, self-care: content translated into English<br>Other links to supportive e-info, if applicable | Comments on Internet Search<br>- Sequence of sites popping up<br>- Major (official) eHealth site<br>- Links to other supportive sources |
|---|---|---|
| | • In Austria, home remedies are commonly used, and SNI is also proposed among the home remedies for bronchitis.<br>[Notably, Germany has a higher ABC (+14%) than Austria. Despite these differences, both countries were given the score 2 for correlation analysis: in both countries, insurance companies, clinicians, hospitals, and naturopathic medicine sites most often included and/or proposed SNI first-line, in line with their popular tradition of "Nasenpflege". In the sensitivity analysis, Austria was augmented in score and given a score of 2.5. Germany, as having "no" official link to a SNI recommendation on the main e-site by the Health Authority, was given a score of 1.5.] | |
| **Italy**<br>↓<br>↓<br>↓<br>↓<br>↓<br>↓<br>↓<br>**Medicines 'to relieve'**<br>↓<br>**Decongestants first-line** for stuffy nose<br>↓<br>↓<br>Saline gargling for 'some'<br>↓<br>AB message | https://www.issalute.it/index.php/la-salute-dalla-a-alla-z-menu/r/raffreddore?highlight=WyJyYWZmcmVkZG9yZSJd<br>Raffredorre (common cold) :<br>• Introduzione : ...//...<br>• Sintomi: ...//....<br>• Cause:...//...<br>• **Terapia:**<br>- Colds generally resolve in 5-10 days. Therefore it is possible to manage the symptoms by following some simple general rules, without consulting your doctor. It may be useful to drink plenty of fluids to replenish those lost due to increased sweating and runny nose; help yourself restore your energy also through a healthy diet, low in fat, high in fibre, with fresh fruit and vegetables, but without forcing yourself to eat, since with a cold, you can lose your appetite.<br>- To help relieve the symptoms, you can use **some medicines** available in pharmacies without a prescription or **remedies such as:** painkillers and antipyretics such as paracetamol and ibuprofen which can reduce fever and also act as painkillers; **decongestants for stuffy nose. These medications are generally safe for older children and adults, but may not be suitable for infants, young childre**n, pregnant women, people with chronic illnesses, or who take certain other medications. It is therefore advisable to always read the information leaflet that accompanies the medicine and follow the instructions for the recommended dosage.<br>- "Some people find relief from sore throats and stuffy noses by gargling with salt water and menthol gum." In newborns and young children, nasal rinses with saline solution (salt water) are very useful. In contrast, antihistamines, cough syrups or **antibiotics** (**effective only against bacteria while colds are caused by viruses**) (read the hoax), complementary treatments and alternative medicine, such as echinacea and herbal medicines, are usually not treatments recommended for colds as there is no evidence on their effectiveness, and they can cause unpleasant side effects. | ISSalute.it = the eHealth portal hosted by Instituto Superiore di Sanita= Official Italian National Institute of Health (ISS), the main centre for research. It is though, a different site than under its own name, retrieving other documents than under its name.<br>https://www.issalute.it/index.php/la-salute-dalla-a-alla-z-menu/a/analgesici - introduzione<br>ISSalute.it does not recommend SNI for cold, but rather paracetamol and ibuprofen first line;<br>gargling with salt water and strangely menthol gum are proposed at end of the e-site just for 'some' people that may find relief, apart from children who generally benefit. The connotation created by the addition of 'menthol gum' for sore throats and stuffy noses reduces the credibility in SNII. The site then recognises last line that in newborns and young children, nasal rinses with saline solution (salt water) are very useful. The site does not communicate convincingly. The statement at the end gets lost discrediting too many options at a same time: antihistamines, cough syrups or **antibiotics (effective only against bacteria while colds are caused by viruses**), the complementary and alternative medicine, such as echinacea and herbal medicines, are usually not treatments recommended for colds as there is no evidence on their effectiveness, and they can cause unpleasant side effects." |
| | **Qualitative research summary**<br>Gargling with saline SNI was found to be a doubtful recommendation, saying "some may benefit" for relief from sore throats and stuffy noses, so little convincing and – by its oral administration – rather addressing laryngitis: the option is proposed last-line on the platform, after emphasis on relief by several medicating options. The same paragraph positioned ABs for bacterial infection, thereby possibly strengthening patients' misbeliefs for the need of an AB, as he/she cannot distinguish viral or bacterial infections. | |
| Luxembourg<br><br>AB message +++ | The site 'Santé.Public.lu (= Santé.Secu.lu in 2025) does not recommend SNI for flue or bronchiolitis on its main pages, also not in children. The only message that pops up is: Cold or flue : no antibiotiques!<br>https://santesecu.public.lu/fr/publications/f/feuille-coloriage-antibiotiques-fr-de.html | SNI is not traditional in Luxembourg.<br>Santé.lu is he official eHealth portal of Direction de la Santé, Luxembourg -> There is no straight forward page to 'rhume'. |

| **Country +** sequence elements from algorithm | **Relevant retrieved sections & Qualitative Research Summary**<br>- Structure of the e-site<br>- Section Treatment, self-care: content translated into English<br>Other links to supportive e-info, if applicable | **Comments on Internet Search**<br>- Sequence of sites popping up<br>- Major (official) eHealth site<br>- Links to other supportive sources |
|---|---|---|
| <br><br><br><br><br><br><br>Flyer 2012<br><br><br><br>AB-message<br><br>Self-treatment<br><br><br><br>Saline | Yet, disease mongering and vaccination is done under the page of "la grippe' (the flue) popping up when searching for the word 'common cold'.<br>*https://sante.public.lu/fr/espace-citoyen/dossiers-thematiques/g/grippe-saisonniere.html*<br>*https://sante.public.lu/fr/espace-citoyen/dossiers-thematiques/g/grippe-saisonniere/grippe.html*<br>https://santesecu.public.lu/fr/campagnes/2023/infections-respiratoires-covid-grippe-pneumo-bronchiolite.html<br><br>Yet, a small flyer for consumers (2012) was identified upon in-depth screening and opening 'all' documents possible: saline was recommended between the lines:<br>https://sante.public.lu/fr/publications/b/bloc-note-medecins-antibio-conseils-fr-de.html<br>In case of cold, cold or flu, do not use antibiotics<br>…//…<br>General tips (flyer 2012, not in current flyer 2025):<br>• Rest and make sure your living spaces are well ventilated.<br>• Drink plenty of water, herbal teas and juices rich in vitamins and without added sugars.<br>• Make sure you eat a balanced and varied diet (rich in vitamins, fruits and vegetables).<br>• If you have a cough or cold, u**se an inhaler or an isotonic salt water nasal spray to to clear your airways.**<br>• To relieve sore throats and avoid the feeling of dry throat, you can use an antiseptic and analgesic throat spray, or suck on small ice cubes. Adults and older children can also suck antiseptic and analgesic throat lozenges.<br>• In case of fever and pain, antipyretic and analgesic medications can be used.<br>relieve you.<br><br>Yet, this saline recommendation was not sustained systematically in subsequent years. A new site has been developed [Accessed 21.02.2025]: no longer recommendation of saline<br>https://santesecu.public.lu/dam-assets/fr/publications/r/rhume-grippe-pas-antibio-fr-de-pt/rhume-grippe-pas-antibiotiques-fr.pdf | The site does not recommend SNI for flue or bronchiolitis on its main pages, also not in children, yet does neither strongly promote other medicines, except for antivirals and or vaccination under the flue. Yet, a small flyer for consumers against ABC we incidentally opened mentions tips, such as i**nhaler or an isotonic salt water nasal spray to clear your airways** between the lines, as well as antiseptic/analgesic throat sprays and suckling ice cubes in case of a sore throat and antivirals for flue in patients at risk, and therefore received score 0.5 in the sensitivity analysis. |
| | ***Qualitative Research Summary - Luxembourg***<br>Luxembourg had a much simpler health portal (Portail Sante.lu, changed in 2025 to SanteSecu.lu [13].<br>• It currently has no information on 'rhume': the previous e-site did not recommend SNI for the common cold or bronchi(oli)tis in children.<br>• Yet, the site of this country did neither propose medicines, as was found in the other countries, except for antiseptic/analgesic throat sprays and suckling ice cubes in case of a sore throat, and antivirals for the flue in patients at risk (see Inventory Table S2).<br>Moreover, after opening and screening all relevant links possible, a flyer from 2012 with the objective to reduce ABC was found, recommending SNI between the lines for relief of common colds [14]. To note, this is no longer the case in 2025: the new brochure for "rhume or grippe?" fully focuses on ABs, yet proposing as solution only hand hygiene, using a handkerchief, and to use of ABs for a bacterial infection (no saline mentioned) [15]. This country therefore received a score of 0.5 (rather than score 0) in the sensitivity analysis. | • |
| Netherlands | Thuisarts.nl<br>https://www.thuisarts.nl/verkouden/ik-ben-verkouden<br>Ik ben verkouden = "I have a cold"<br>• Video | • Clearcut first-line recommendation of saline, also steaming, while decongestants are 2$^{nd\text{-}line}$ with an immediate negative connotation by Thuisarts.nl. |

| **Country +** sequence elements from algorithm | **Relevant retrieved sections & Qualitative Research Summary**<br>- Structure of the e-site<br>- Section Treatment, self-care: content translated into English<br>Other links to supportive e-info, if applicable | **Comments on Internet Search**<br>- Sequence of sites popping up<br>- Major (official) eHealth site<br>- Links to other supportive sources |
|---|---|---|
| Salt water (saline) first-line<br><br>Decongestant 2nd line, limited use<br>↓<br>↓<br>↓<br>↓<br><br><br>AB message | [eHealth site follows the same recommendations]<br>Content from the video:<br>- Viral cause explained<br>- Clear message in video:<br>• **Medicines are not necessary for a cold.**<br>[do you suffer from a stuffy nose?]<br>You can use n**asal drops or nasal spray:** You can use 3x a day:<br>• **Saltwater: spray or drops**<br>• Xylometazoline drops: **a medicine not to be used for longer than 1 week.** If you use them for too long, they can damage the mucous membranes.<br>• Both without a prescription at the pharmacy or drugstore<br>• Steaming is also possible, but does not shorten the duration....<br>A cold will go away on its own.<br>SOMETIMES IT TAKES A LONG TIME.<br>You may continue to cough, sneeze and sniffle for a few weeks<br><br>ANTIBIOTICS<br>• do not help against a cold.<br>• Also NOT if the cold lasts longer.<br>• Also NOT if the snot changes color (text specifies: for example yellower or greener). **Even then, medications are of no use.**<br>• Colds are caused by a virus and antibiotics do not help against that.<br>When to call a doctor?<br>• If you become short of breath: You notice that you are breathing difficult or quickly.<br>• If you become increasingly ill<br>• Have a fever above 38° for more than 5 days | • The recommendations are clearly consistently followed by several other platforms that pop up, such as:<br>• Arts en zorg .nl<br>• KNO: Association Nose Throat Ear specialists<br>• ZEL: first-line self-management = non-profit advice and support organization for primary care<br>• Gezondheidsplein.nl<br>= from Solvo.nl<br>= eHealth platform of 5000 active specialists in health care |
| | ***Qualitative analysis summary - Netherlands***<br>The first hit for 'common cold' went directly to the relevant page of the governmentally monitored eHealth sites, Thuisarts.nl.<br>Strikingly persuasive simple cues, explaining the symptoms and causes and offering SNI first-line before medications such as nasal decongestant sprays or drops: the latter medicines were discouraged by a clear-cut short message on both e-sites.<br>The official internet site also provides instructions on how to make saline solution for nasal irrigation.<br>There seems also consistency throughout other guidelines. | • |
| **Norway**<br><br><br><br><br>AB message<br>↓<br>Medicine first-line: decongestants & | https://www.helsenorge.no/sykdom/ore-nese-hals/forkjolelse/<br>Forkjølelse -> 21 hits<br>Treatment<br>• There is no effective treatment for the common cold, but it goes away on its own within a few days. However, there are drugs that can alleviate some of the symptoms.<br>• Antibiotics do not help against viruses. Incorrect use of antibiotics increases antibiotic resistance in society.<br>Medicines<br>• Medicines used for colds are pain relievers, decongestants and cough medicines. One should be cautious with the use of medicines for children under six years of age. | Helse Norge<br>Does not recommend saline, rather decongestants<br><br><br>In Norway, 700,000 people are now dependent on nasal sprays (https://www.newsendip.com/in-norway-700000-people-are-addicted-to-nasal-sprays/ ) Newsendip, March 5, 2024<br><br>You still can find some places recommending 'Neseskylling kan lindre' = nasal irrigation can help, such as by NEL (="Norsk Helse |

| **Country +** sequence elements from algorithm | **Relevant retrieved sections & Qualitative Research Summary**<br>- Structure of the e-site<br>- Section Treatment, self-care: content translated into English<br>Other links to supportive e-info, if applicable | **Comments on Internet Search**<br>- Sequence of sites popping up<br>- Major (official) eHealth site<br>- Links to other supportive sources |
|---|---|---|
| cough medicines, painkillers, mucosal decongestants<br>↓<br>↓<br>↓<br>Self-treatment<br><br><br><br>No saline<br>↓<br>↓<br>↓<br>↓<br>↓<br>↓<br>↓<br>↓<br>↓<br>↓<br>↓<br>↓<br>↓<br>↓<br>↓<br>↓<br>↓<br>↓<br>↓<br><br>**Unless in children:**<br>Salt water drops<br>↓<br>↓<br>↓<br>↓<br>↓<br>Yet also offering the option of decongestants | • Painkillers can be used for a sore throat, muscle pain or headache. Paracetamol and ibuprofen can be bought without a prescription. Paracetamol in excessive doses can cause serious liver damage. Check the package insert to avoid overdose.<br>• **Mucosal decongestants can open a stuffy nose for several hours. They should not be used for more than a week.**<br>• Cough medicines are a collective term for several different substances. Documentation of the effect is uncertain. Cough suppressants may contain morphine-like substances. These work by inhibiting the cough center in the brain. They should not be used when driving.<br>**Self-help**<br>• **When you have a cold, you should not overexert yourself. Make sure you drink enough fluids and stay warm.**<br>• **Some people try to inhale steam to reduce the swelling of the nasal mucosa. A warm bath or shower can clear up the nose.**<br>Prognosis<br>• The common cold usually goes away on its own within 7-10 days. However, the cough can last for three weeks or more.<br>• It is usually not necessary to see a doctor for a cold, but it is recommended if you are not better after two weeks or if your general condition is greatly reduced. This applies especially to the very youngest and oldest.<br>• Sometimes an additional infection occurs with a cold. It could be an ear, lung or sinus infection. Pneumonia can be serious, especially for the elderly, young children and those with chronic diseases. (underlined = links to these indications)<br>• Pneumonia = linked bacteria & antibiotics [although the cause says: Lung inflammation (pneumonia) is usually caused by an infection with bacteria or viruses. The infection causes inflammation in the lungs, making it harder to breathe]<br><br>In children:<br>https://www.helsenorge.no/sykdom/lunger-og-luftveier/tett-nese-hos-barn/<br>Tett nese hos barn = Congested nose in children<br>Measures to keep the nose as open as possible can prevent many ailments. Effective swallowing and the sense of smell depend on an open nose. If the nose is blocked, the heating, filtering and moistening of the air is reduced. A stuffy nose can also disturb sleep at night.<br>**Effect of nasal drops**<br>There are two main types of nasal drops. Both are available both as drops in single-dose containers and as a spray. It is most comfortable for the child that the nasal drops are at room temperature.<br>• Salt water drops<br>**Physiological saline drops contain sodium chloride and are used in the same strength by adults and children. Saline drops can be used to rinse away or remove mucus and crusts in the nose. In many children, this may be enough to relieve nasal congestion.**<br>• Breast milk ...<br>• Mucosal decongestant nasal drops contain xylometazoline or oxymetazoline and have different strengths for children in different age groups.<br>• Mucous decongestant nasal drops prevent the mucous membrane in the nose from swelling by causing the blood vessels in the mucosa to contract. Less mucus is then also formed.<br>**It is not certainly documented that these medicines have an effect on children.** | informatikk AS") Norwegian Health informatics AS , also in Alternative medicin (Skandinavisk yoga og meditasjonsskole) |

<table>
<tr><th>Country + sequence elements from algorithm</th><th>Relevant retrieved sections & Qualitative Research Summary<br>- Structure of the e-site<br>- Section Treatment, self-care: content translated into English<br>Other links to supportive e-info, if applicable</th><th>Comments on Internet Search<br>- Sequence of sites popping up<br>- Major (official) eHealth site<br>- Links to other supportive sources</th></tr>
<tr><td></td><td>Used over a long period of time, nasal drops can lead to chronic nasal congestion. It is therefore important that they have the right strength based on the child's age, that you follow the recommended dosage and that the nasal drops are not used for more than a couple of days in a row.</td><td></td></tr>
<tr><td></td><td>Qualitative Research Summary<br>The official eHealth site rather proposes medicines to relieve common colds first-line, with a clearcut first-line recommendation for decongestants.<br>The layout of the eHealth site moreover - likely unwillingly - leads to a discrediting perception about saline drops for use in children (unless this would be a translation error in our analysis): the site puts a sentence on nasal saline under the item “nasal drops” saying: “It is not documented with certainty that these medicines have an effect in children.”<br>If parents are not visiting the more specific children-oriented page where saline is proposed [16], this sentence is misleading because saline is not a medicine: consumers counselling the site may not make the difference between saline and medicinal drops, as both are available OTC in the pharmacy.<br>As like Denmark, this country proposes and emphasizes decongestants for symptom relief first-line, it is given the score of 1.</td><td></td></tr>
<tr><td>Spain<br><br>Page popping up for ‘flue’ = ‘gripe’<br><br>↓<br>↓<br>↓<br>↓<br>↓<br>↓<br>↓<br><br>“Medications” - antivirals<br>↓<br>↓<br>↓<br>↓<br>↓<br>↓<br>↓<br>↓<br>↓<br>↓<br>↓<br>↓<br>↓<br><br>AB message<br>↓</td><td>https://informaciongripea.sanidad.gob.es/preguntas-y-respuestas-mas-frecuentes.html<br>Blue banner on top: “La prevención es la mechor medida” (Prevention is the best measure)<br>Qué es la gripe? What’s the flu?<br>Very long page: https://informaciongripea.sanidad.gob.es/preguntas-y-respuestas-mas-frecuentes.html#informacion6<br>• INFORMACIÓN GENERAL: …//…<br>• TRANSMISIÓN: …//…<br>• PREVENCIÓN: ..//..<br>• SÍNTOMAS: ..//..<br>• GRUPOS ESPECÍFICOS: ..//..<br>• DIAGNÓSTICO : ..//..<br>• TRATAMIENTO:<br>Are there medications to treat the flu?<br>• Yes. The new flu virus is treated like the seasonal flu, with medications that relieve the symptoms: to reduce fever, cough, muscle pain…<br>• In Spain, antiviral treatments are also available that may be useful: oseltamivir (Tamiflu®) and zanamivir (Relenza®). None of these medications cure. They act by reducing the duration of the disease by approximately one day and reduce the possibility of complications. For all these reasons, they are currently only indicated in people with the flu who require hospitalization or those who, until now, have a higher probability of suffering complications.<br>• In these people, it is considered that, when diagnosed with flue, the benefit of the administration of these drugs outweighs the risk of possible adverse effects that may be present in any medication.<br>• Like all medications, Tamiflu and Relenza have side effects and contraindications, therefore, they should always be prescribed by medical professionals. The irresponsible use of these medications can contribute to creating virus resistance, which would reduce their effectiveness.<br>• Currently, Spain has a reserve of antivirals of more than 15 million treatments, which are already being used in cases where it is indicated. These medications are not dispensed in pharmacies.<br>• The flu is caused by a virus, so antibiotics are not indicated. They are only useful, always under medical prescription, in the event of bacterial complications.</td><td>Links for common cold were screened, but hits are meddled by links going to platforms from all over the world, including e.g. Mexico, Columbia, Chili, Peru, Pan American Health Organisation, and also many sites originating in English from the United States (e.g. Mayo CLINIC and CDC, popping up with their Spanish translations. There is also a clear dilution with other indications such as flue , or respiratory infection in general. Adding “portal de salud” was added, identifying the “Ministerio de Sanidad” : 115 hits related to salina, 118 hits related to “resfriado” (many linked to programmes against antibiotics), but as a consumer, no acceptable speed for identification of guidelines possible.<br><br>When looking for “sanidad” .es, rather Sanidad.es pops up, before the governmental site: this is a commercial site reviewing various options in various articles on the site; also old (non-proven) remedies are reviewed, one referring to ‘antibiotic power’ of onion (fuelling the idea antibiotics work), yet not SNI. The owner is unclear (‘SOBRE NOSOTROS’ is unlinked)<br><br>Apart from many other commercial sites, offering medicines for a cold, the site of the Spanish paediatric association and Formation of paediatricians (AEP= was identified and analysed. https://enfamilia.aeped.es/temas-salud/catarro-resfriado: overall, the messages propose various medicines and discredit saline as not being proven effective, and even propose saline as a decongestant.<br><br>If any, steam and saline, the first option in Netherlands – well explained how to do - appear last line.</td></tr>
</table>

| **Country +** sequence elements from algorithm | **Relevant retrieved sections & Qualitative Research Summary**<br>- Structure of the e-site<br>- Section Treatment, self-care: content translated into English<br>Other links to supportive e-info, if applicable | **Comments on Internet Search**<br>- Sequence of sites popping up<br>- Major (official) eHealth site<br>- Links to other supportive sources |
|---|---|---|
| Vaccination<br>↓<br>↓<br>↓<br>↓<br>↓<br>↓<br>↓<br>↓<br>↓<br>↓ | • **VACUNA:**<br>- Is there an authorized vaccine for the new virus?<br>- What influenza A vaccine should I get?<br>- How many doses of vaccine do I need?<br>- What side effects does the vaccine have?<br>- How useful is the vaccine?<br>- Who has to get vaccinated?<br>- When is vaccination going to start in our country?<br>- For which people is vaccination against the new pandemic virus (H1N1) 2009 indicated?<br>- How many vaccines have been ordered?<br>- Will there be safe vaccines for children with egg allergies? And for adults allergic to eggs?<br>- How is this vaccine similar to the seasonal flu vaccine?<br>[extensive explanation on vaccines] | |
| Page on 'resfriado'<br><br>Medicines proposed in a box<br>↓<br>↓<br>↓<br>↓<br>↓<br>↓<br>↓<br>↓<br>↓<br>Zinc<br><br>Onion infusion<br><br>Gingher<br><br>Eucalyptus<br><br>Chicken soup<br><br>**No saline**<br><br><br><br>AB-message: no | When searching for "resfriado" on Sanidad.es: the first message is:<br>https://www.sanidad.es/deshazte-del-resfriado/<br>**How to get rid of a cold in 24 hours:**<br>[Medicamentos] (= Medicines)<br>It hasn't happened to you that you wake up one morning with a feeling of heaviness and with your nasal passages a little blocked. Generally, it is a flu or cold. This condition is very common.<br>In many cases this illness lasts between three and five days. You feel so bad, broken down and muscularly sore, that you don't want to do anything. But the reality is that at work they will not give you permission to be absent.<br>So the best thing is to take action at the first symptoms of a cold. It is not about "stunning" it, the ideal is to do a quick cure. Here we present some options, they work in 24 hours.<br>The first thing you should keep in mind is that if symptoms persist, you should consult a specialist doctor. These tips are simply tips to make your life easier.<br>You can also read: 5 curiosities about sleep that you may not know<br>Natural tips to cure a cold:<br>- Zinc: (picture of various pills): When the cold starts, take a zinc or vitamin C capsule to stop the symptoms before they appear more strongly.<br>- Onion infusion: The antibiotic power of garlic helps protect the body's immune system. Garlic tea can help you before the cold progresses<br>- Ginger with lemon : favours mucus expulsion<br>- Eucalyptus inhalation : Helps stop throat irritation. As it is steam, it enters the respiratory tract and decongests them. 10 grams of leaves in a cup of boiling water is enough.<br>- Chicken soup: The grandmothers always gave us a good chicken soup for everyone. It's not a coincidence. So having a cup when you first feel the symptoms is important to relieve them.<br>- A warm water bath: Warm water takes away muscle pain. The heat also opens the bronchi, which will make the disease leave your body quickly.<br>[No messages on antibiotics on the many articles we opened, related to 'resfriados]<br>To note: searching the site for "salina", SNI was only found under the topic of sinusitis and bronchitis in children.<br><br>When re-searching for a link with messages on antibiotic use by the governmental site [10/08/2024], it was deviated to the following link : | |

| **Country +** sequence elements from algorithm | **Relevant retrieved sections & Qualitative Research Summary**<br>- Structure of the e-site<br>- Section Treatment, self-care: content translated into English<br>Other links to supportive e-info, if applicable | **Comments on Internet Search**<br>- Sequence of sites popping up<br>- Major (official) eHealth site<br>- Links to other supportive sources |
|---|---|---|
| AB-message searched for: | https://resistenciaantibioticos.es/es/lineas-de-accion/comunicacion/campanas/campana-pran-2023-antibioticos-protegernos-es-su-trabajo-el-tuyo-usarlos-bien<br>"Antibióticos, protegernos es su trabajo. El tuyo, usarlos bien"<br>*"Antibiotics, protecting us is their job. Yours, using them well"*<br>The main objective of this campaign is to raise awareness among the entire population about the proper use of antibiotics. Another year, we employ a touch of humor to explain that using antibiotics for "a job they are NOT made for", i.e. for a flu, a headache or a cold, causes them to then stop working when less we need, which is against bacteria. And it is that if we misuse antibiotics, we are endangering the health of everyone: that of people, that of animals, but also that of the environment.<br><br>This plan seems to be implemented separately by:<br>Agencia Española de Medicamentos y Productos Sanitarios (AEMPS)<br><br>The plan seems to be implemented independently of the complaints:<br>https://www.resistenciaantibioticos.es/es/lineas-de-accion/comunicacion/campanas | |
| | ***Qualitative Research Summary - Spain***<br>In Spain (the country with highest ABC in our analysis), hits were meddled by links going to platforms from all over the world, including e.g. Mexico, Columbia, Chili, Peru, Pan American Health Organisation, and to many sites in English originating from the United States (e.g. Mayo CLINIC and CDC), popping up with their Spanish translations.<br>• There is also a clear dilution with other indications such as flue, or respiratory infections in general.<br>• Adding the target word "portal de salud" identified the Ministerio de Sanidad y Politica Social (Sanidad.gob.es) with 118 hits related to "resfriado": the opening of the links deviated the screen to other indications such as the flue, or respiratory infections in general, with the recommendation of antivirals and vaccination, but as a consumer, there appeared no acceptable speed for identification of simple clearcut guidelines for the common cold.<br>• A banner tells "prevention is the main measure" [17], while on the flue page, the reference to inappropriate use of ABs is last-line.<br>• The message against AB use is rather covered by a separate agency, the Agencia Española de Medicamentos y Productos Sanitarios (AEMPS) [18].<br>• Even more confusing, the similarly named site "Sanidad.es" brings more focused consumer messages on "How to get rid of a cold in 24 hours" yet not recommending saline [19]. | |
| **Sweden**<br>**1177**<br><br><br><br><br>↓<br>↓<br>↓<br>↓ | https://www.1177.se/sjukdomar--besvar/infektioner/forkylning-och-influensa/<br>Förkylning – 1177 (= common cold)<br>https://www.1177.se/sjukdomar--besvar/infektioner/forkylning-och-influensa/forkylning/#:~:text=Prova%20koksaltl%C3%B6sning%20om%20du%20%C3%A4r,du%20k%C3%B6pa%20receptfritt%20p%C3%A5%20apotek<br>https://www.1177.se/sjukdomar--besvar/infektioner/forkylning-och-influensa/forkylning/<br>• **Symtom :**<br>It often only takes a few days from being infected until you notice the first symptoms of a cold. These are common symptoms:<br>- fatigue<br>- runny nose or nasal congestion<br>- sore throat | 1147.se : official site with information and services in health care by the Swedish authorities:<br>Saline is clearly recommended first-line, even for ear infection (Halsfluss)<br><br>To note: 1177 is the official site with information and services in health and care by the Swedish authorities<br>• "1177 is all of Sweden's gathering place for i. We offer healthcare advice, information, inspiration and e-services. We are available 24 hours a day at 1177.se and on phone 1177 for healthcare advice."<br>• "On 1177.se you can get advice on health and information about diseases and which clinics you can contact. Log in to |

| **Country +** sequence elements from algorithm | **Relevant retrieved sections & Qualitative Research Summary**<br>- Structure of the e-site<br>- Section Treatment, self-care: content translated into English<br>Other links to supportive e-info, if applicable | **Comments on Internet Search**<br>- Sequence of sites popping up<br>- Major (official) eHealth site<br>- Links to other supportive sources |
|---|---|---|
| ↓<br>↓<br>↓<br>↓<br>Self-treatment<br>↓<br>↓<br>↓<br>↓<br>Saline first-line<br>↓<br>↓<br>**Decongestant only if spray does not help**<br>↓<br>↓<br>↓<br>↓<br>↓<br>↓<br>↓<br>↓<br>↓<br>↓<br>↓<br>↓<br>↓<br>↓<br>↓<br>↓<br>↓<br>↓<br>↓<br>↓<br>↓<br>↓ | - cough.<br>A cold can also cause a fever, but it is usually not high. It is more common for children to get a fever with a cold than for adults.<br><br>Trouble in the eyes and ears: Sometimes the cold can cause symptoms from the eyes and ears. The eyes may become watery and sometimes a bit persistent. You may also experience ringing in the ears, and hearing may be impaired. Sometimes you may also get a ringing in the ear or hear a tone.<br><br>The cough may remain for a few weeks The symptoms usually last for a few days. The symptoms you get differ from person to person. You usually feel well again after one to two weeks.<br>The cough may remain for a while after the cold. This is because the mucous membranes are still sensitive. It has usually passed after two to four weeks.<br><br>• **Vad kan jag göra själv? ..//.. = What can I do myself?**<br>Stay home until you feel better if you feel sick.<br>• There are things you can do yourself to ease the discomfort of a cold.<br>• You may need to rest, but that doesn't mean you have to lie down. Often it is good to be up and moving. However, you should not play sports or exercise if you have an infection in your body.<br>**Try saline solution if your nose is blocked**<br>• **First of all, try nasal drops or nasal sprays containing saline solution** if your nose is very congested. You can buy them without a prescription at a pharmacy.<br>You can use the nasal spray for a limited time<br>• You can try decongestant nasal sprays if saline does not help. You can buy it without a prescription at a pharmacy. Use the nasal spray according to the instructions on the package and for a maximum of ten days. Long-term use of nasal sprays can cause nasal congestion that is difficult to get rid of.<br>Tips if you have a sore throat:<br>Hot drinks such as tea can feel soothing if you have a sore throat. Ice cream and yogurt can also feel good. Try out what works best for you.<br>Sometimes it feels better to have something to suck on. You can try throat lozenges available in grocery stores and pharmacies. They do not reduce the swelling in the throat, but it can feel soothing. The best thing for the teeth is to use sugar-free tablets.<br>There are several different types of lozenges and throat sprays that relieve sore throats in different ways. Ask at a pharmacy what can possibly relieve your symptoms.<br>Drinking can help if you have a cough<br>Try drinking more than usual if you have trouble with a cough. Drink makes mucus less sticky and easier to cough up. Avoid being in smoky environments. The smoke irritates the airways and often makes the cough worse. If you smoke yourself, there are many benefits to quitting smoking.<br>There is no research that shows that over-the-counter cough medicines have any clear effect. But some people think they help.<br>Drinking a lot is usually as effective as expectorant drugs.<br>If you have a fever<br>When you have a cold, you can sometimes have a fever that lasts for a few days. It does not necessarily mean that you have contracted a more serious illness. Drink more than usual if you have a fever. The body loses more fluid then.<br>It can also feel nice to be cool in the room where you sleep. | read your medical record and do your care tasks. Call phone number 1177 for healthcare advice around the clock."<br>• 1177 gives you advice when you want to feel better."<br>• "If you are on a visit in Sweden and become ill or get injured, call 1177 for information about illnesses and about where to find your nearest healthcare centre. In case of emergency, call 112 for an ambulance. " |

| **Country +**<br>sequence elements from algorithm | **Relevant retrieved sections & Qualitative Research Summary**<br>- Structure of the e-site<br>- Section Treatment, self-care: content translated into English<br>Other links to supportive e-info, if applicable | **Comments on Internet Search**<br>- Sequence of sites popping up<br>- Major (official) eHealth site<br>- Links to other supportive sources |
|---|---|---|
| ↓<br>↓<br>↓<br>↓<br>↓<br>↓<br>↓<br>↓<br>↓<br>↓<br>↓<br>↓<br>↓<br>↓<br>↓<br>↓<br>AB-message<br>+<br>Explanation | You often do not need to treat a fever when you have a cold. If you sleep poorly or have body aches due to a fever, you can get help from antipyretic drugs. Here you can read more about over-the-counter medicines for pain and fever.<br><u>Sleep with the head elevated</u><br>Try to lie with your head high in bed. It can help if you have a cough and nasal congestion that makes it difficult to sleep. The mucous membranes become less swollen. The lungs also work better in an upright position.<br>You can make a bed with some extra pillows. Place the pillows under the mattress and it will feel more comfortable for the back.<br><u>Home remedies</u><br>There are many home remedies that are claimed to help with a cold. Eating garlic or large amounts of vitamin C is one such home remedy. But there is no research showing that it either helps or prevents colds.<br><u>Natural remedies:</u> In Sweden, there are several herbal remedies that are approved to relieve colds. Some are made from the plant echinacea, which is also called red sun hat. There is no research to show that herbal remedies have any effect when you have a cold, but some people find that they help.<br>• **När och var ska jag söka vård? = When and where should I seek care? ..//..**<br>• **Så smittar förkylning = When and where should I seek care? ..//..**<br>• **Behandling = Treatment**<br>Colds are usually caused by viruses and then treatment with antibiotics does not help. The body's immune system takes care of the healing and the cold goes away on its own.<br><u>What happens in the body?</u><br>The common cold is the most common form of respiratory infection. A cold affects the upper respiratory tract, mainly in the nose.<br>Viruses or bacteria cause the mucous membranes to become infected. They then become swollen and produce more mucus than usual. The mucus can be thin or thick. It can be green or yellow in color.<br>Cold viruses come in many varieties<br>When you catch a cold, you have been infected by a cold virus. Such viruses come in many different variants. After a cold, you have some protection against a new infection of the same virus type. But the risk is high that you will come across new variants of the cold virus.<br>Some get colds more easily than others<br>People are differently susceptible to infection. Some people almost never catch a cold, while others get it several times a year.<br>You who have asthma or other types of chronic lung diseases often get more prolonged colds. This is also the case if you have a weakened immune system.<br><br><u>How can I reduce the risk of colds?</u><br>Your body can be better able to fight off infections if you exercise, have healthy eating habits and get enough sleep.<br>The immune system can be weakened by alcohol. It can also worsen if you have too much stress or too little sleep for a long time.<br>…//…<br><br>Instructions how to prepare saline:<br>• https://www.1177.se/Halland/sjukdomar--besvar/ogon-oron-nasa-och-hals/nasa-och-luktsinne/rensa-nasan-med-koksalt/<br><br>Consistent lay press messages: | |

| **Country +** sequence elements from algorithm | **Relevant retrieved sections & Qualitative Research Summary**<br>- Structure of the e-site<br>- Section Treatment, self-care: content translated into English<br>Other links to supportive e-info, if applicable | **Comments on Internet Search**<br>- Sequence of sites popping up<br>- Major (official) eHealth site<br>- Links to other supportive sources |
|---|---|---|
| | https://www.meds.se/tipsochrad/bli-av-med-forkylning-snabbare-tips/#:~:text=Att%20sk%C3%B6lja%20n%C3%A4san%20med%20koksaltl%C3%B6sning,slemmet%20som%20kan%20vidbeh%C3%A5lla%20infektion)<br><br>https://www.lakemedelsverket.se/sv/om-lakemedelsverket/press-och-nyheter/kort-om/sa-behandlar-du-din-forkylning-pa-basta-satt#hmainbody1) | |
| | ***Qualitative Research Summary - Sweden***<br>For Sweden, the first hit for 'common cold' went directly to the relevant page of the governmentally monitored eHealth site 1177.se.<br>Overall, persuasive simple cues, explaining the symptoms and causes and offering SNI first-line before medications such as nasal decongestant sprays or drops: the latter medicines were discouraged by a clear-cut short message on both e-sites.<br>Internet site also provides instructions on how to make saline solution for nasal irrigation. Country is given the maximum score of 3. | |

4 Belgische Commissie voor de coördinatie van het antibioticabeleid (BAPCOC). :. Missie. https://overlegorganen.gezondheid.belgie.be/nl/advies-en-overlegorgaan/commissies/belgische-commissie-voor-de-coordinatie-van-het-antibioticabeleid#:~:text=De%20Belgische%20Commissie%20voor%20de,de%20bestrijding%20van%20antimicrobi%C3%ABle%20resistentie.

5 Agence nationale de sécurité du médicament et des produits de santé (ANSM). Plan hivernal" [Winter plan]: https://ansm.sante.fr/dossiers-thematiques/plan-hivernal [Acces: 24.06.2024]

6 VIDAL. ABC of medicines. Rhume (rhinite). Mise à jour : 08 décembre 2023. https://www.vidal.fr/maladies/nez-gorge-oreilles/rhume-rhinite.html [Last access: 02.08.2025]

7 Agence nationale de sécurité du médicament et des produits de santé (ANSM). En cas de rhume, évitez les médicaments vasoconstricteurs par voie orale ! [In case of a cold, avoid oral vasoconstrictor medications!https://ansm.sante.fr/actualites/en-cas-de-rhume-evitez-les-medicaments-vasoconstricteurs-par-voie-orale [Acces: 24.06.2024]

8 Agence nationale de sécurité du médicament et des produits de santé (ANSM). Le virus respiratoire syncytial (VRS) [Respiratory Syncytial Virus]. https://ansm.sante.fr/dossiers-thematiques/le-virus-respiratoire-syncytial-vrs [Acces: 24.06.2024, 02.08.2025]

[9] Agence nationale de sécurité du médicament et des produits de santé (ANSM). Rhume, nez qui coule, nez bouché ? Attention : l'utilisation des vasoconstricteurs expose à des risques, soyez vigilants! [Cold, runny nose, blocked nose? Warning: the use of vasoconstrictors exposes you to risks, be vigilant!] https://ansm.sante.fr/actualites/rhume-nez-qui-coule-nez-bouche-attention-lutilisation-des-vasoconstricteurs-expose-a-des-risques-soyez-vigilants

[10] Santé.fr. Rhinopharyngite ou Rhume. Information proposée par Antibio'Malin [Rhinopharyngoitis or Common cold. Information proposed by Antibio'Malin] https://www.sante.fr/antibiomalin-pour-savoir-comment-bien-utiliser-les-antibiotiques/rhinopharyngite-ou-rhume [Acces: 10.08.2024]

[11] Santé.fr. Rhume. Information proposée par Service Public d'Information en Santé [Common cold. Information by the Public Health Information Service]. https://www.sante.fr/rhume-rhinite [Acces: 10.08.2024]

[12] Österreichische Sozialversicherung. Erkältung: Was kann ich selbst tun? [Common cold: what can I do myself?] https://www.sozialversicherung.at/cdscontent/?contentid=10007.862784&portal=svporta

[13] Santé.lu (=Santé.secu.lu: https://santesecu.public.lu/fr.html). Flyer 2010. Cold or flue : no antibiotiques! https://santesecu.public.lu/fr/publications/f/feuille-coloriage-antibiotiques-fr-de.html

[14] Santé.lu (=Santé.secu.lu). Flyer 2012: Bloc-note médecins "Conseils pour une guérison rapide lors d'une infection virale (rhume grippe,…)" https://sante.public.lu/fr/publications/b/bloc-note-medecins-antibio-conseils-fr-de.html

[15] Santé.lu (=Santé.secu.lu). Brochure. Rhume ou grippe ? Pas d'antibiotiques ! Cold or or flu? No antibiotics. https://santesecu.public.lu/dam-assets/fr/publications/r/rhume-grippe-pas-antibio-fr-de-pt/rhume-grippe-pas-antibiotiques-fr.pdf

[16] Helse Norge. Tett nese hos barn [= Congested nose in children]. https://www.helsenorge.no/sykdom/lunger-og-luftveier/tett-nese-hos-barn/ [Access: 27.06.2024]

[17] Sanidad.gob.es. Ministerio de Sanidad y Politica Social. https://informaciongripea.sanidad.gob.es/preguntas-y-respuestas-mas-frecuentes.html#informacion6 & Qué es la gripe? [What's the flu?] https://informaciongripea.sanidad.gob.es/preguntas-y-respuestas-mas-frecuentes.html#informacion6

[18] Agencia Española de Medicamentos y Productos Sanitarios (AEMPS). Antibióticos, protegernos es su trabajo. El tuyo, usarlos bien. [Antibiotics, protecting us is their job. Yours, using them well] https://resistenciaantibioticos.es/es/lineas-de-accion/comunicacion/campanas/campana-pran-2023-antibioticos-protegernos-es-su-trabajo-el-tuyo-usarlos-bien [10/08/2024]

[19] Sanidad.es. Resfriado. Deshazte del resfriado en 24 horas. [The common cold: How to get rid of a cold in 24 hours.] https://www.sanidad.es/deshazte-del-resfriado/ [10/08/2024]

**Table S3. Scoring** of the eHealth communication for the Common Cold of 13 Countries from the Inventory (for scores: see main manuscript)

**Scoring system**: item analysed: SNI, decongestants, medicines (antivirals), vaccination (if applicable), complication

(1) countries with scores 2-3: clear SNI recommendation: score 3 refers to clearcut first-line integration of SNI in the recommendations for treatment, irrespective of former traditional use, while score 2 refers to broadly established SNI traditional use, yet not first-line support for relief on the official site, yet by (link to) e-sites by official associations, health care workers and/or other communications

(2) countries with score 1: SNI traditional, but other medicine, such as decongestants, being recommended first-line (no support for SNI, unless by separate link for children)

(3) countries with score 0: SNI not traditional, overall emphasis on medicine, sometimes vaccination and complications; no (or no clearcut) saline recommendation

For the sensitivity analyses of the correlation, the scores were simplified using only 3 scores (score 0-1-2), and vice versa, were finetuned for relevant small differences in recommendations by using additional half scores if mixed recommendations were found (saline between the lines/at end of page) (Table S4).

| | | eHealth recommendation of SNI for the common cold | | | | No traditional use | | | | Category (see Figure 1 &Table 2 for categories) |
|---|---|---|---|---|---|---|---|---|---|---|
| | | Authority: SNI first-line in communication? | SNI +/- steaming traditional? | Supported on e-sites by health care workers and other communications? | Decongestant recommended first line **(No or no clearcut saline )** | Saline message lost (between the lines, or only at end of the page? | Major focus on other medication (even if negative advice) | e-Site emphasis on Vaccination | Complications (disease mongering) | |
| 1 | Austria | no | **yes** | **yes** [ample support] | no | | | | | 2 |
| 2 | Belgium | no | no | no (only children) | no | **yes** (end of page) | yes (decongestants) | - | - | 0 |
| 3 | Denmark | no | **yes** | - | **yes** | **no** | yes | - | - | 1 |
| 4 | Estonia | no | **yes** | (yes: one source identified) | **yes** | yes | mixed | - | - | 1 |
| 5 | Finland | no | **yes** | **yes** (also by video how to prepare) [ample support] | (limit use) | | | | | 2 |
| 6 | France | no | no | no (only children) | (limit use) + other | yes end of page + VIDAL) | yes | Yes | Yes | 0 |
| 7 | Germany | no | **Yes** | **yes** [ample support] | (short-term) | | | | | 2 |
| 8 | Italy | no | no | no (only children) | **yes** | yes (gargling helping 'some') | no | no | - | 0 |
| 9 | Luxembourg | no | no | - | no | limited (1 flyer) | no | Yes | - | 0 |
| 10 | Netherlands | **yes (also video)** | no** | **yes** | (limit use) | | | | | 3 |
| 11 | Norway | no | yes | - | **yes** | (Separate section on children) | - | - | - | 1 |
| 12 | Spain | no | no | - | no | **no** | **yes (antivirals)** (other remedies – no saline) | Yes | - | 0 |
| 13 | Sweden | **yes** | yes | - | no (if saline does not help) | | | | | 3 |
| | | yes = maximum Score 3 | yes = score 2 if traditional SNI *AND* amply supported in communications by other health care providers, and general media | | yes = score 1 *AND* traditional SNI (no general e-support) | Emphasis on medicines, vaccination and complications No (or no clearcut) saline recommendation SNI not traditional for common cold | | | | |

- : not readily identified by internet searches

* France : first-line in VIDAL, but not elsewhere, and perceived to be lost by the overloading other information with emphasis on nasal sprays, medicines and complications needing medicines

** Netherlands : SNI is not traditional, yet proposed since start of the eHealth site (2012 - not known if inserted at launch in 2009)

- not observed, + observed but in balance with the rest of the site, and ++ excessive, overwhelming, also on internet

Mixed = when one major eHealth site recommends/overemphasizes medicine first-line (SNI only between the lines), but other important clinicians' recommendations don't and propose SNI first-line.

Table S4. **Sensitivity Analyses:** Data used from ECDC and the Score Category used in the Main Analysis and Sensitivity Analyses

| S4-A. Community sector | DDD 2022 J01 | Category Main analysis | Category Sensitivity analysis (simplified) | Category Sensitivity analysis (finetuned) | Reason for changing score |
|---|---|---|---|---|---|
| Austria | 8.8 | 2 | 2 | 2.5 | Overt official support first-line by official insurance company |
| Belgium | 19.1 | 0 | 0 | 0.5 | One one site: saline recommended at end of e-page |
| Denmark | 13.33 | 1 | 1 | 1 | Unchanged: traditional saline, but decongestant first-line: no saline found |
| Estonia | 10.78 | 1 | 1 | 1.5 | Overt official support first-line in brochure by health care organisations |
| Finland | 10.5 | 2 | 2 | 2 | Unchanged : general support on site & market |
| France | 22.56 | 0 | 0 | 0 | Unchanged: 'medicalised' lost between other medical options and complications |
| Germany | 10.03 | 2 | 2 | 1.5 | Decreased score, as no official info on saline, despite high popularity of Nasenpflege & ample independent support by health care workers |
| Italy | 20.05 | 0 | 0 | 0 | Unchanged: not traditional, decongestant first-line: |
| Luxembourg | 17.64 | 0 | 0 | 0.5 | One flyer: saline between the lines |
| Netherlands | 8.32 | 3 | 2 | 3 | Unchanged : clear recommendation for saline |
| Norway | 14.02 | 1 | 1 | 1.5 | Traditional saline, but decongestant first-line: yet saline found for children under separate heading |
| Spain | 21.70 | 0 | 0 | 0 | Unchanged : nowhere saline recommended |
| Sweden | 9.6 | 3 | 2 | 3 | Unchanged : clear recommendation for saline |

| Analysis | Pearson | | | Spearman | | |
|---|---|---|---|---|---|---|
| | Correlation coefficient | *p*-value | 95% CI | Correlation coefficient | *p*-value | 95% CI |
| Main | *r* = -0.9049 | 0.000001 | [-0.9715, -0.7057] | *r*s = -0.9450 | 0.000001 | [-0.9895,-0.7379] |
| Simplified categories | *r* = -0.9446 | 0.000001 | [-0.9836, -0.8207] | *r*s = -0.9376 | 0.000002 | [-0.9880,-0.7078] |
| Finetuned | *r* = -0.9421 | 0.000001 | [-0.9829, -0.8132] | *r*s =-0.9267 | < 0.00001 | [-0.9782, -0.7676] |

https://www.statskingdom.com/correlation-calculator.ht

| S4-B. Community sector | DDD 2019 J01 | DDD 2022 J01 | DDD 2023 J01 (community sector) | Category Main analysis | Category Sensitivity analysis (finetuned) |
|---|---|---|---|---|---|
| Austria | 9.77 | 8.8 | 9.5 | 2 | 2.5 |
| Belgium | 19.78 | 19.1 | 19.1 | 0 | 0.5 |
| Denmark | 13.44 | 13.33 | 14.3 | 1 | 1 |
| Estonia | 10.24 | 10.78 | 11.2 | 1 | 1.5 |
| Finland | 12.56 | 10.5 | 11.1 | 2 | 2 |
| France | 23.34 | 22.56 | 22.4 | 0 | 0 |
| Germany | 11.37 | 10.03 | 11.7 | 2 | 1.5 |
| Italy | 19.80 | 20.05 | 21.2 | 0 | 0 |
| Luxembourg | 19.75 | 17.64 | 18.7 | 0 | 0.5 |
| Netherlands | 8.68 | 8.32 | 8.8 | 3 | 3 |
| Norway | 13.61 | 14.02 | 14.2 | 1 | 1.5 |
| Spain | 23.27 | 21.70 | 22.5 | 0 | 0 |
| Sweden | 10.33 | 9.6 | 8.7 | 3 | 3 |

2023 data: Bindel LJ, Seifert R. Most European countries will miss EU targets on antibacterial use by 2030: historical analysis of European and OECD countries, comparison of community and hospital sectors and forecast to 2040. *Naunyn Schmiedebergs Arch Pharmacol*. 2025;398(8):10195-10220. doi:10.1007/s00210-025-03887-5

| Analysis | Spearman | | |
|---|---|---|---|
| | Correlation coefficient | *p*-value | 95% CI |
| Main | *r*s = -0.9422 | < 0.000001 | [-0.9889, -0.7262] |
| Finetuned | *r*s =-0.9862 | < 0.00001 | [-0.9975, -0.9268] |

| S4-C<br>Hospital sector | DDD 2023<br>J01 (community sector) | Category<br>Main analysis | Category<br>Sensitivity analysis<br>(finetuned) |
|---|---|---|---|
| Austria | 1.8 | 2 | 2.5 |
| Belgium | 1.5 | 0 | 0.5 |
| Denmark | 1.9 | 1 | 1 |
| Estonia | 1.5 | 1 | 1.5 |
| Finland | 1.8 | 2 | 2 |
| France | 1.7 | 0 | 0 |
| Germany | - | 2 | 1.5 |
| Italy | 1.9 | 0 | 0 |
| Luxembourg | 1.5 | 0 | 0.5 |
| Netherlands | 0.8 | 3 | 3 |
| Norway | 1.3 | 1 | 1.5 |
| Spain | 1.6 | 0 | 0 |
| Sweden | - | 3 | 3 |
| | No significant correlation | $r_s$ = -0.1322<br>$p$ = .698 | $r_s$ = -0.3037<br>$p$ = .364 |

2023 data: Bindel LJ, Seifert R. Most European countries will miss EU targets on antibacterial use by 2030: historical analysis of European and OECD countries, comparison of community and hospital sectors and forecast to 2040. *Naunyn Schmiedebergs Arch Pharmacol*. 2025;398(8):10195-10220. doi:10.1007/s00210-025-03887-5

**Table 5. Propositions of eHealth communication aspects to be investigated in the future and to integrate in OneHealth programmes, as to reduce ABC and antimicrobial resistance, and as to benefit populations world-wide.**

| **I. Proposed investigations in eHealth communication:** |
|---|
| **Specific on ABs:** |
| • **Patient perceptions and attitudes in response to AB messages**: find out their attitudes to see a doctor and their perception/expectations that the message creates for needing an AB: if the different messages are scorable differently, correlate with the ABC in the model; see also empowerment of message by proposing SNI<br>• **Delay in AB prescribing**: number of days or weeks at which the eHealth portal refers the reader to seek a doctor's advice for the common cold and other relevant indication; also analyse the number and type of conditions for referral, as to detect a potential counterproductive effect of listing these in driving patients to seek a doctor for an AB |
| **Specific to SNI:** |
| • **Change-behaviour attitudes to proposing SNI for the cold**: empowerment of the AB message by proposing SNI, if presented just for hygiene/nose cleansing/rinsing, versus also for its other benefits in the common cold, and which of these are the most motivating to induce a switch: effective in 'relieving' nasal congestion, soothing throat aches and post-nasal drip associated cough, resolving faster the cold (reducing the cold duration), reducing the ABC, safe for self-care…<br>• **Spill-over effect to other indications**: Expand analysis with focus on SNI to other indications (e.g. sinusitis, sore throat, cough, ear infection...): similarly scoring possible & correlate with ABs?<br>• **Impact of instructions how to prepare & availability OTC on attitudes**. |
| **Other eHealth treatment recommendations:** |
| • **Role of restrictive messages**: study of the potential counterproductive impact of (too high focus on) restrictive use of decongestants on patient's attitudes and needs for seeking doctor's advice and so ABs. What if not improving beyond the allowed period of use?<br>• **Role of other non-pharmacological measures:** impact of recommending steaming, but also hygiene interventions such as hand washing, distancing…. : if first-line proposed, possibly perceived as trivial to change-behaviour: so, where and how to communicate them at best [they are to be followed for more serious infections]. |
| **Structure e-site**: |
| • **Impact of a well-structured persuasive communication versus complex or confusing e-messages**: analyse the elements of persuasive communication itself (e.g. starting from sites in countries with the lowest ABC, Netherlands and Sweden) and investigate the perceptions and attitudes directly with readers/patients: e.g.<br>- Accessibility of the site |

- Logic and wording of the messages
- Motivational elements that engage into following the recommendations
- The public audience that is attracted/influenced by the message

**II. Possible integration in One Health programmes - clinically oriented towards outcomes in the long-term:**

- **Doctors' and public education (guidelines, flyers, other promotion raising awareness):**
  - Integrate persuasively SNI in guidelines for primary care and flyers for the common cold
  - Evaluate the impact of implementing the local guideline/campaign with doctors in primary care and with the public
- **In parallel: if not available, develop (or adapt) a well-structured, trustful common cold page on the eHealth site,** based on countries with low ABC (e.g. the Netherlands) – easy ,navigation, mobile accessible
- **Evaluate the impact of implementing the e-Site**, also including the inexpensive accessible option of saline nasal drops or irrigation:
  - in a non-traditional test market with high ABC, as a test case: e.g. Luxembourg
  - in a traditional test market, currently promoting decongestant medicines first-line (e.g. Norway, Denmark); monitor the outcome on ABC
- **For countries with more fragmented communications**: simplify to a well-trusted and easily accessible eHealth site, ensuring the website's name and logo are consistent, with focus on clear, simple and reliable ccommunications, and user-friendly navigation, as in the Netherlands
- **Overall, strengthen the local public e-site**, making sure the site is attractive, and accessible by the Public while ensuring so-called search-enzyme- optimization and raising awareness about the s-site
- **Long-term : Policy development : Study (in)consistency** across other professional and other guidelines: e.g. e-messages by clinicians' associations, e-promotion (pharmacy, druggists, supermarkets and e-sale platforms) and other influencers on the internet.
- **Monitor access (click-through rate):** consider the success, so impact of promotional e-sites versus the official public e-site
